# Germline Variants Influence Chronic Liver Disease Progression through Distinct Pathways

**DOI:** 10.1101/2025.09.16.25335186

**Authors:** Marijana Vujkovic, David E Kaplan, Jonas Ghouse, Bao-Li Loza, Joseph Brancale, Adam Lewis, David Y Zhang, Michael G Levin, Olivia J Veatch, Josephine P Johnson, Carolin V Schneider, Anurag Verma, Kirk J Wangensteen, Eleonora Scorletti, Dipender Gill, Chigoziri Konkwo, Alexis M Garófalo, Lindsay A Guare, Tae-Wi Schwantes-An, Marco V Abreu, Helene Gellert-Kristensen, Ole B Pedersen, Christian Erikstrup, Johan S Bundgaard, Erik Sørensen, Sisse R Ostrowski, Henning Bundgaard, Kyung Min Lee, Abraham Shaked, Kim M Olthoff, Maarouf A Hoteit, Elizabeth K Speliotes, Yanhua Chen, Antonino Oliveri, Lishi Yin, Luca VC Valenti, Francesco Malvestiti, Daniele Marchelli, Lorenzo Miano, Quentin M Anstee, Ann K Daly, Heather J Cordell, Rebecca Darlay, Niek Verweij, George Hindy, Adam Locke, Kentaro Matsuura, Sumeet K Asrani, Giuliano Testa, Luca A Lotta, Marcus B Jones, Daniel R Dochtermann, Trina M Norden-Krichmar, Craig C Teerlink, Poornima Devineni, Saiju Pyarajan, Daniel J Rader, Yasuhito Tanaka, Benjamin F Voight, Silvia Vilarinho, Lisa A Bastarache, Stefan Stender, Philip S Tsao, Penn Medicine Biobank, DBDS Genomic Consortium, BioVU Biobank, Michigan Genomics Initiative, Regeneron Genetics Center, Indiana Biobank, All Of Us Research Program, Milano Biobank, LITMUS Consortium, VA Million Veteran Program, Timothy R Morgan, Julie A Lynch, Kyong-Mi Chang

## Abstract

Cirrhosis and hepatocellular carcinoma (HCC) are long-term complications of chronic liver disease (CLD). In this large multi-ancestry genome-wide association study of all-cause cirrhosis (35,481 cases, 2.36M controls) and HCC (6,680 cases, 1.76M controls), we identified 27 loci associated with cirrhosis (10 novel) and 11 with HCC (three novel). Three novel cirrhosis loci were replicated in independent cohorts (e.g. *FGF21, RPTOR*, and *IFNL3/4)*. Fifteen cirrhosis loci exhibited differential effects on cirrhosis risk via underlying etiologies, and six HCC loci influenced HCC risk indirectly via cirrhosis. In a gene-burden analysis of rare variants from whole-genome sequencing data in the VA Million Veteran Program (n=102,677), we identified *GSTA5* as a novel cirrhosis-associated gene, while *APOB* and *ATP9B* were associated with and replicated for HCC. A high genetic risk score for cirrhosis was associated with a nearly doubled risk of CLD progressing to cirrhosis (HR=1.94, P=2×10^−68^) and of cirrhosis progressing to HCC (HR=1.65, P=7×10^−08^). Finally, among individuals with chronic hepatitis C who underwent antiviral therapy, cirrhosis risk was modified by variants in *PNPLA3, IFNL3/4*, and *CD81* following pegylated interferon-α therapy, and by *APOE* lead variant following direct-acting antiviral therapy. These findings provide new insights into the complex genetic architecture of CLD progression with potential clinical and therapeutic implications.

## INTRODUCTION

Chronic liver disease (CLD) is a global health concern due to its potential for progression to cirrhosis and hepatocellular carcinoma (HCC) with significant morbidity and mortality. CLD arises from multiple causes including hepatotropic viruses (hepatitis B and C), alcohol, and metabolic dysfunction-associated steatotic liver disease (MASLD) and steatohepatitis (MASH), as well as various heritable and autoimmune conditions. In patients with CLD, chronic hepatocellular injury with inflammation, oxidative stress, and stellate cell activation can promote fibrogenesis and hepatocellular turnover, ultimately leading to liver cirrhosis and HCC.^1-3^ Recent advances in human genetics have improved our understanding of genetic susceptibility to CLD and its progression to cirrhosis and HCC.^4-14^ Two large-scale studies identified 15 cirrhosis-associated and nine HCC-associated variants, and showed that a genetic risk score (GRS) derived from cirrhosis-associated variants predicted cirrhosis onset in individuals with MASLD, as well as HCC in individuals with pre-existing cirrhosis.^6,15,16^ Despite these advances, it remains unclear whether distinct genetic variants drive CLD progression to cirrhosis and/or HCC directly or via specific underlying causes of CLD, such as alcohol, viral, and/or metabolic pathways. Furthermore, the clinical utility of genetic risk prediction for cirrhosis and HCC remains uncertain. This knowledge gap impedes the development of targeted therapeutic strategies and integrated patient risk stratification strategies for preventing and managing liver-related complications.

Here, we leveraged summary-level genetic data from multiple studies and biobanks, including the Million Veteran Program (MVP), to perform the largest multi-ancestry genome-wide and immunogenetic association analysis of cirrhosis and HCC to date. We complemented this with coding variant and gene-burden analyses using whole-genome sequencing (WGS) data from 102,677 MVP participants. Leveraging two decades of electronic health records in MVP, we performed causal mediation analysis of underlying etiologies, multi-state survival modeling of genetic risk scores, developed integrative prediction models of clinical and genetic risk factors, and assessed impact of cirrhosis-associated variants on cirrhosis progression following antiviral therapy in patients with chronic hepatitis C. Finally, we applied drug target Mendelian randomization to prioritize therapeutic targets for cirrhosis and HCC. An overview of all analyses is shown in Figure 1. These findings provide opportunities for Precision Medicine, while advancing our insight to the complex genetic architecture in CLD progression to cirrhosis and HCC.

**Figure 1:**
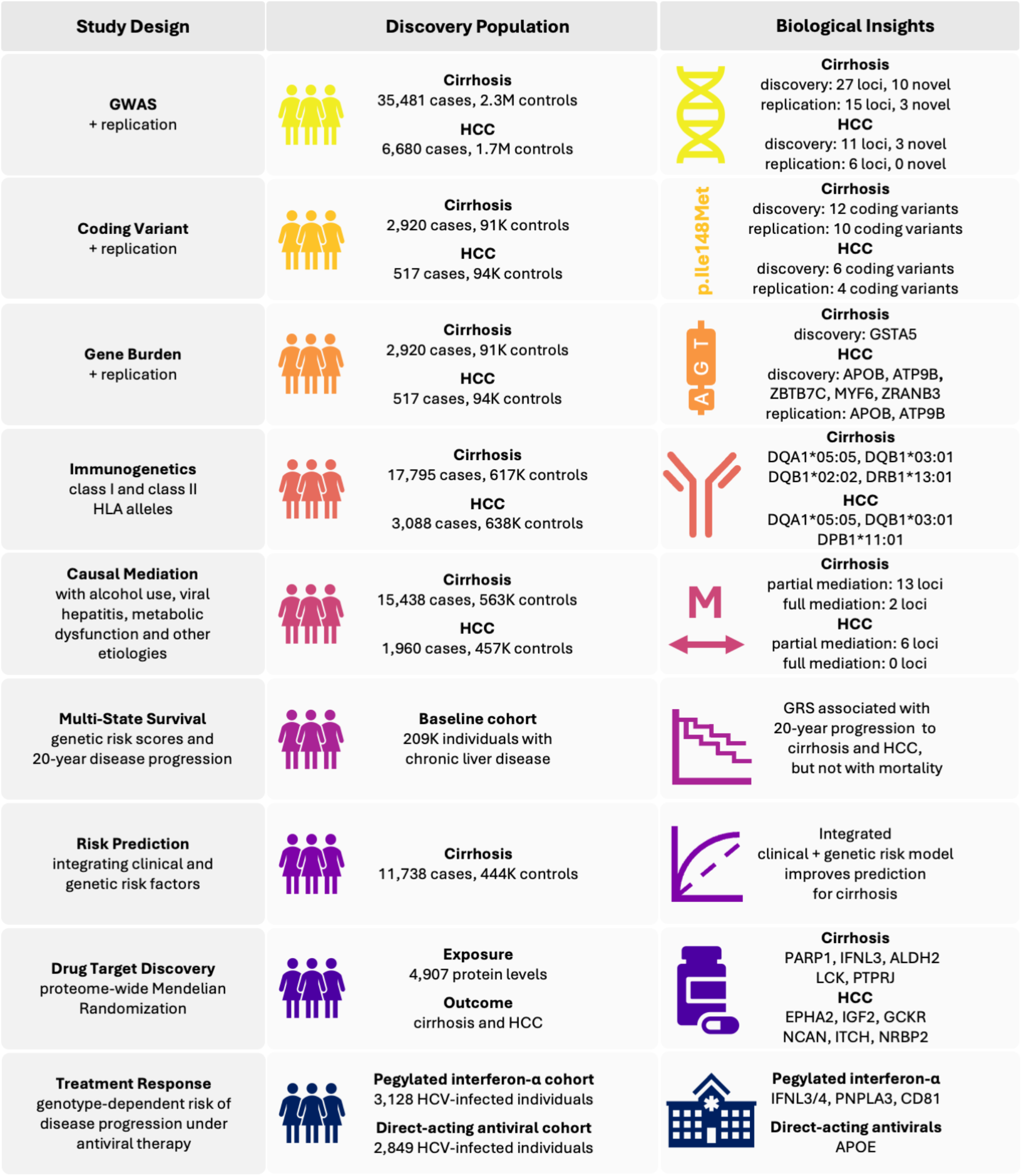
Overview of study design and analyses. Nine complementary analyses were performed in the current study, namely: a common variant GWAS, coding variant analysis, gene-burden analysis, immunogenetic discovery of HLA alleles, causal mediation analysis of underlying etiologies, multi-state survival analysis of genetic risk scores, integrated clinical and genetic risk prediction, drug target Mendelian randomization, and genotype-specific risk of disease progression under antiviral therapy.

## RESULTS

### Comprehensive multi-ancestry discovery of common, coding, and immunogenetic variants for cirrhosis and HCC

We performed multi-ancestry genome-wide meta-analyses of all-cause cirrhosis and HCC, integrating summary statistics from the MVP with those from other cohorts (Figure 2, Table 1). The cirrhosis GWAS included 35,481 cases and 2,359,621 population controls, and the HCC GWAS included 6,680 HCC cases and 1,756,387 controls. Most participants were of European descent (85% for cirrhosis, 81% for HCC), with smaller contributions from East-Asian (7% for cirrhosis, 9% for HCC), African American (5% for cirrhosis, 7% for HCC), and Hispanic American (2% for cirrhosis, 3% for HCC) populations (Supplementary Table 1).

**Table 1.** Common variant discovery and replication for cirrhosis

| Gene | Novel | Chr | Position (hg38) | EA | NEA | Multi-Ancestry Discovery |  |  |  | Replication Cohort |  |  |  |
| --- | --- | --- | --- | --- | --- | --- | --- | --- | --- | --- | --- | --- | --- |
|  |  |  |  |  |  | EAF | Beta | SE | P-value | EAF | Beta | SE | P-value |
| *PDE4B* | - | 1 | 65969060 | A | G | 0.49 | 0.05 | 0.01 | $2.7 \times 10^{-08}$ | 0.47 | 0.04 | 0.02 | $0.011^{-08}$ |
| *MTARC1* | - | 1 | 220796686 | G | A | 0.73 | 0.10 | 0.01 | $1.1 \times 10^{-27}$ | 0.73 | 0.16 | 0.02 | $3.2 \times 10^{-18}$ |
| *PDK1* * | Yes | 2 | 173420650 | A | T | 0.16 | 0.17 | 0.03 | $5.9 \times 10^{-08}$ | 0.41 | -0.11 | 0.09 | $0.242^{-08}$ |
| *PPARGC1A* * | Yes | 4 | 30482764 | G | C | 0.20 | 0.00 | 0.01 | $0.729^{-08}$ | 0.19 | -0.04 | 0.04 | $0.313^{-08}$ |
| *HSD17B13* | - | 4 | 87310713 | T | C | 0.26 | -0.13 | 0.01 | $1.4 \times 10^{-42}$ | 0.23 | -0.14 | 0.02 | $3.7 \times 10^{-12}$ |
| *ADH1B* | - | 4 | 99318162 | C | T | 0.66 | 0.18 | 0.02 | $3.5 \times 10^{-16}$ | 0.94 | 0.21 | 0.04 | $6.8 \times 10^{-08}$ |
| *CXXC4*† | Yes | 4 | 104494029 | A | G | 0.40 | -0.05 | 0.01 | $2.1 \times 10^{-07}$ | 0.38 | -0.04 | 0.02 | $0.033^{-08}$ |
| *HFE* | - | 6 | 25917997 | C | T | 0.06 | 0.14 | 0.02 | $8.4 \times 10^{-15}$ | 0.05 | 0.03 | 0.04 | $0.405^{-08}$ |
| *HLA* | - | 6 | 32685100 | G | T | 0.26 | -0.13 | 0.01 | $1.4 \times 10^{-26}$ | 0.29 | -0.03 | 0.03 | $0.381^{-08}$ |
| *TRIB1* | - | 8 | 125487187 | G | C | 0.71 | -0.07 | 0.01 | $2.9 \times 10^{-17}$ | 0.71 | -0.11 | 0.02 | $1.7 \times 10^{-09}$ |
| *TOR1B* | - | 9 | 129804387 | A | G | 0.10 | -0.10 | 0.02 | $1.7 \times 10^{-10}$ | 0.08 | -0.19 | 0.06 | $6.8 \times 10^{-04}$ |
| *HKDC1* | - | 10 | 69224180 | C | G | 0.61 | -0.07 | 0.01 | $5.2 \times 10^{-14}$ | 0.59 | -0.10 | 0.02 | $4.6 \times 10^{-04}$ |
| *GPAM* | - | 10 | 112162067 | A | G | 0.71 | -0.07 | 0.01 | $1.8 \times 10^{-14}$ | 0.71 | -0.12 | 0.02 | $1.3 \times 10^{-11}$ |
| *CD81* * | Yes | 11 | 2370014 | T | C | 0.26 | -0.10 | 0.02 | $1.2 \times 10^{-07}$ | 0.11 | -0.10 | 0.04 | $0.012^{-08}$ |
| *BDNF* | Yes | 11 | 27672444 | G | C | 0.70 | -0.05 | 0.01 | $3.8 \times 10^{-09}$ | 0.70 | -0.03 | 0.02 | $0.060^{-08}$ |
| *LXRA* | Yes | 11 | 48148165 | A | G | 0.72 | 0.06 | 0.01 | $2.4 \times 10^{-09}$ | 0.73 | -0.02 | 0.04 | $0.665^{-08}$ |
| *SH2B3* | - | 12 | 110929440 | T | C | 0.19 | -0.28 | 0.04 | $4.8 \times 10^{-14}$ | 0.02 | -0.14 | 0.17 | $0.422^{-08}$ |
| *ALDH2* | - | 12 | 111730205 | A | G | 0.25 | -0.28 | 0.03 | $1.3 \times 10^{-16}$ | 0.01 | 0.07 | 0.22 | $0.761^{-08}$ |
| *SERPINA1* | - | 14 | 94378610 | T | C | 0.02 | 0.45 | 0.03 | $4.6 \times 10^{-60}$ | 0.01 | 0.72 | 0.05 | $4.6 \times 10^{-39}$ |
| *ACE* | Yes | 17 | 63256498 | A | G | 0.18 | -0.07 | 0.01 | $6.3 \times 10^{-09}$ | 0.18 | -0.02 | 0.02 | $0.400^{-08}$ |
| *RPTOR* | Yes | 17 | 80320965 | C | G | 0.05 | -0.12 | 0.02 | $3.8 \times 10^{-09}$ | 0.05 | -0.13 | 0.04 | $0.001^{-08}$ |
| *TM6SF2* | - | 19 | 19277691 | T | A | 0.09 | 0.24 | 0.01 | $1.6 \times 10^{-78}$ | 0.08 | 0.28 | 0.03 | $3.1 \times 10^{-25}$ |
| *IFNL3/4* | Yes | 19 | 39248489 | G | C | 0.20 | 0.09 | 0.01 | $1.7 \times 10^{-22}$ | 0.21 | 0.07 | 0.02 | $2.0 \times 10^{-04}$ |
| *APOE* | - | 19 | 44912921 | T | G | 0.26 | -0.09 | 0.01 | $4.7 \times 10^{-21}$ | 0.24 | -0.12 | 0.02 | $2.3 \times 10^{-10}$ |
| *FGF21* | Yes | 19 | 48758111 | C | T | 0.65 | -0.05 | 0.01 | $4.4 \times 10^{-08}$ | 0.68 | -0.07 | 0.02 | $6.0 \times 10^{-05}$ |
| *MBOAT7* | - | 19 | 54173307 | G | C | 0.59 | -0.05 | 0.01 | $1.2 \times 10^{-09}$ | 0.57 | -0.05 | 0.02 | $0.002^{-08}$ |
| *PNPLA3* | - | 22 | 43928847 | G | C | 0.26 | 0.39 | 0.01 | $8.1 \times 10^{-446}$ | 0.24 | 0.50 | 0.02 | $8.5 \times 10^{-179}$ |

\* Genome-wide signal in African American Discovery cohort ( $P < 5.0 \times 10^{-08}$ , Supplemental Table 4)
† Genome-wide signal in European Discovery cohort ( $P < 5.0 \times 10^{-08}$ , Supplemental Table 4)
Chr = chromosome; EA = effect allele; NEA = non-effect allele; EAF = effect allele frequency; SE = standard error

**Figure 2:**
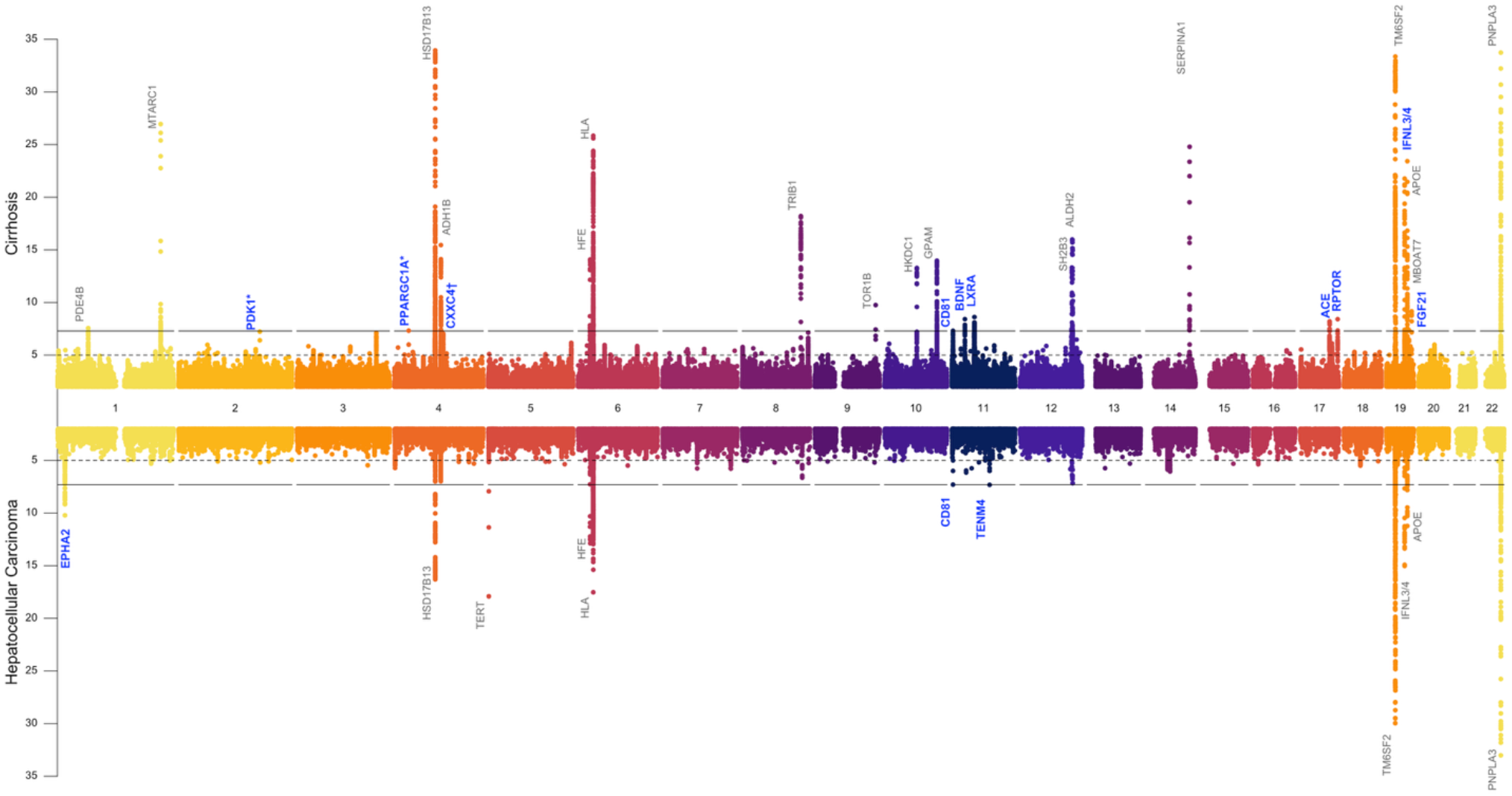
Genome-wide association results for cirrhosis and hepatocellular carcinoma. Two Manhattan plots display genome-wide association statistics for cirrhosis (top) and hepatocellular carcinoma (HCC; bottom). Each point represents a single nucleotide polymorphism (SNP), plotted by genomic position on the x-axis and –log_10_(P-value) on the y-axis. The HCC panel is inverted to facilitate visual comparison. Multi-ancestry association signals are shown, with ancestry-specific genome-wide significant associations (P < 5×10^−8^) overlaid for completeness. These include *PDK1* and *PPARGC1A* (African American) and *CXXC4* (European) for cirrhosis, and *CD81* (African American) for HCC. Select genome-wide significant loci are annotated with the gene in each region most likely to be functionally relevant, with blue font indicating novel loci. The gray dashed lines indicate the genome-wide significance threshold. This figure highlights both shared and ancestry-specific genetic architecture for cirrhosis and HCC.

Cirrhosis and HCC case definitions were based on ICD9/10 diagnosis codes in most cohorts (Supplementary Table 1 and 2). MVP was the single largest contributor of cases for discovery, providing 50% of cirrhosis cases and 46% of HCC cases, where we further refined the phenotypes to improve specificity by requiring national cancer registry confirmation for HCC and restricting cirrhosis cases to those with a high FIB-4 score (>2.67) at the time of diagnosis (Supplementary Table 3). The cirrhosis cases in MVP encompassed a heterogeneous mix of etiologies for CLD, with a high prevalence of alcohol-related liver disease (57%), chronic hepatitis C (43%) and metabolic dysfunction with elevated liver enzymes (55%) which frequently co-occurred.

In MVP GWAS data, we performed discovery analyses of common and immunogenetic variants, and, using recently released WGS data from 102,677 MVP participants, we additionally conducted putative causal coding variant analysis as well as collapsed gene-burden tests of deleterious rare variants.

### Twenty-seven genetic loci are associated with cirrhosis risk

Our all-cause cirrhosis meta-analysis revealed 27 genome-wide significant loci for cirrhosis (P < 5.0×10^−8^ Figure 2, Table 1). Namely, 17 loci were significant in the European-ancestry analysis, three additional loci in African-ancestry analysis, and seven emerged only in the multi-ancestry meta-analysis (Supplementary Table 4). Ten of 27 SNPs had not been previously reported for cirrhosis, including variants near or within *CXXC4, IFNL3*/*4, FGF21, PDK1, PPARGC1A, CD81, BDNF, LXRA, ACE* and *RPTOR*.

### Use of diseased instead of population controls attenuates effects at three cirrhosis loci

In a separate disease progression GWAS conducted within the MVP cohort (n=191,491), we used individuals with CLD (n=191,491) as controls instead of population-based controls. The CLD controls included individuals diagnosed with alcohol-related liver disease, chronic viral hepatitis, metabolic dysfunction with elevated liver enzymes (Met-ALT), and other causes of liver disease based on diagnoses codes as listed in Supplementary Table 2. In this analysis, six loci remained genome-wide significant in the multi-ancestry analysis. Among European Americans, genetic effect estimates were similar for 22 of 25 common loci (ancestry-specific MAF > 1%) between the two control groups, and for all tested loci in African Americans and Hispanics (Supplementary Figure 1). Notably, the effects of *PNPLA3, ADH1B* and *HFE* on cirrhosis risk were significantly attenuated when using CLD controls among Europeans, suggesting that associations at these loci are at least partly driven by susceptibility to underlying risk factors, such as Met-ALT and alcohol use for *PNPLA3*, alcohol use for ADH1B, and hemochromatosis for HFE.

### Replication of twelve known and three novel cirrhosis loci

To replicate common variants associated with cirrhosis, we analyzed data from 8,806 independent cirrhosis cases and 328,061 population controls, including the Michigan Genomics Initiative^17^, LITMUS Consortium^18^, Milano Biobank^19^, Regeneron Genetics Center^20^, and All Of US Research Program^15,21^(Supplementary Table 5). Twenty-three loci showed concordance in direction of effect between the discovery and replication cohort, which is unlikely to have occurred by chance (Table 1, two-tailed binomial P < 0.001). Fifteen loci reached Bonferroni-corrected statistical significance (P < 0.0019), of which three were novel (*FGF21, IFNL3/4*, and *RPTOR)*. Nominal replication was observed for two novel loci, *CD81* and *CXXC4* (P < 0.05).

### Identification of eleven HCC loci with effect attenuation in cirrhosis-restricted controls

The HCC GWAS with population controls identified 11 genome-wide significant loci, including three novel loci: *EPHA2, CD81*, and *TENM4* (Figure 2, Table 2). In a separate analysis within the MVP cohort using only cirrhotic patients as controls, no loci remained genome-wide significant (Supplementary Table 6). For nine of 11 HCC loci, effect estimates were significantly attenuated towards the null in European Americans, with a comparable pattern of effect attenuation among African and Hispanic Americans (Supplementary Figure 2). These findings suggest that using population controls for HCC may capture not only direct effects on HCC risk but also variant effects mediated through susceptibility to CLD and cirrhosis, which could lead to larger genetic effect estimates compared to analyses restricted to disease-specific controls.

**Table 2.**
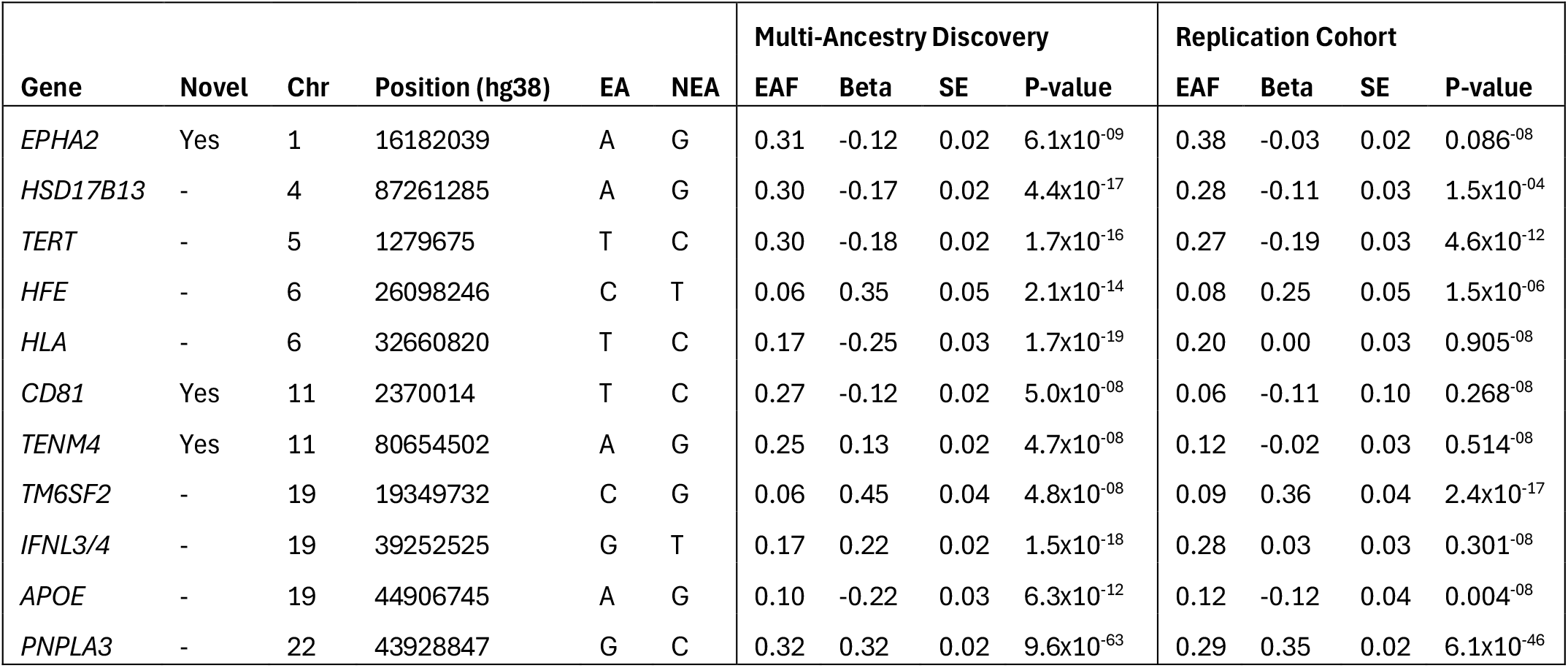
Common variant discovery and replication for HCC

### Six HCC-associated loci were replicated

To replicate common variants associated with HCC, we analyzed data from a cohort of nine independent studies encompassing a total of 4,995 HCC cases and 706,432 population controls, including the Michigan Genomics Initiative, Penn Transplant Cohort, Milano Biobank, European ALD-HCC Study, Japanese HCV-HCC Study, Indiana Biobank, BioVU Biobank, Korea Biobank, Copenhagen Hospital Biobank and Danish Blood Donor Study, and Regeneron Genetics Center (Supplementary Table 7).^9,16,17,19,20,22-26^ Ten of 11 HCC loci had direction of effects in the replication cohort consistent with the direction in the discovery cohort, which is unlikely to have occurred by chance (Table 1, two-tailed binomial P = 0.006). Six SNPs achieved replication at the Bonferroni-corrected significance level, including *PNPLA3, TM6SF2, TERT, HFE, HSD17B13*, and *APOE* (P<0.004).

### Coding variant analysis replicates ten putative causal variants for cirrhosis and four for HCC

Three lead variants were missense in the cirrhosis GWAS (*PNPLA3* p.Ile148Met, *MTARC1* p.Thr165Ala, and *FGF21* p.Leu174Pro), whereas *PNPLA3* p.Ile148Met was the only coding lead variant in the HCC GWAS. All lead coding variant were replicated in the common variant replication cohort (Table 1 and 2). To identify additional putative causal variants for cirrhosis, we evaluated an additional eight missense and one splice-site variant in high linkage with non-coding lead variants in the Regeneron Genetics Center exome sequencing dataset (D′ > 0.95 and r^2^ > 0.4, Supplementary Table 8). Seven of nine splice-site and/or coding variants replicated at Bonferroni significance, namely *TM6SF2* p.Glu167Lys, *IFNL3* p.Lys70Arg, *HSD17B13* c.812+2dup, *ADH1B* p.His48Arg, *GPAM* p.Ile43Val, *SERPINA1* p.Glu366Lys, and *APOE* p.Cys130Arg (P<0.006). For HCC, we evaluated five missense variants in high linkage with non-coding lead variants, three of which replicated at a Bonferroni-corrected significance level (P<0.01, *TM6SF2* p.Glu167Lys, *IFNL3* p.Lys70Arg, and *HSD17B13* c.812+2dup). In summary, we replicated 10 coding variants (e.g. 3 common and 7 rare) for cirrhosis and four for HCC (e.g. 1 common and 3 rare).

### Five HLA class II alleles linked to cirrhosis and/or HCC in immunogenetic analysis

We performed a dedicated immunogenetic discovery analysis of the major histocompatibility complex (MHC) region on chromosome 6p21.3 in MVP, using imputed four-digit class I and II HLA alleles, while adjusting for age, sex, and common etiologies of liver disease. We identified several class II HLA alleles (but no HLA class I alleles) associated with cirrhosis risk at a Bonferroni-corrected P-value threshold of 0.0002 (Supplementary Table 9). Specifically, we observed protective associations for HLA-DQA1*05:05 and HLA-DQB1*03:01 with cirrhosis in both European and African American ancestry groups, whereas HLA-DQB1*02:02 in European and HLA-DRB1*1301 in African Americans were additionally associated with increased risk of cirrhosis. Immunogenetic analysis of HCC showed that the two class II alleles protective against cirrhosis (HLA DQA1*05:05 and DQB1*03:01) were also protective against HCC in European and African Americans. In addition, HLA-DPB1*11:01 was associated with increased HCC risk in Hispanic American individuals (Supplementary Table 9).

### Gene-burden analysis identiXies one novel gene associated with cirrhosis and six with HCC

We next investigated if aggregated rare coding variants are associated with cirrhosis (1,971 European cases vs 68,603 controls; 949 African cases vs 22,505 controls) and HCC (326 European cases vs 70,744 controls; 191 African vs 23,624 controls) using WGS data from MVP. In gene-burden tests of rare protein-truncating variants (predicted loss-of-function, pLoF) and deleterious missense variants (inferred by AlphaMissense), we identified *GSTA5* as being associated with cirrhosis among African Americans (Supplementary Table 10). The association at *GSTA5* was primarily driven by a high-confidence stop-gained variant (p.Trp155Ter, β = 1.059, SE = 0.25, P = 2.7×10^−05^). This pLoF variant occurs at a population frequency of ∼0.6% in individuals of African ancestry but is essentially monomorphic in non-African populations. In the Regeneron sequencing dataset, *GSTA5* p.Trp155Ter showed a consistent direction of effect but did not reach statistical significance (P = 0.133, Supplementary Table 10). For HCC, gene-burden analyses identified *APOB, DMBT1, ATP9B*, and *ZBTB7C* among European Americans, and *MYF6* and *ZRANB3* among African Americans (Supplementary Table 10).

### Replication conXirms APOB and ATP9B as novel genetic contributors to HCC

In a combined gene-burden meta-analysis across six external sequencing cohorts including the Milano Biobank, Regeneron Genetics Center, All of Us Research Program, UK Biobank, and Mass General Brigham Biobank, *APOB* and *ATP9B* replicated at Bonferroni significance for HCC, while *ZBTB7C* showed a nominal association (Supplementary Table 10). For cirrhosis, *GSTA5* gene burden showed a suggestive association with a consistent direction of effect, though it did not reach statistical significance (P = 0.08).

### Fifteen loci impact cirrhosis risk through underlying causes of liver disease

Given that many cirrhosis-associated variants are associated with alcohol use disorder and MASLD, we investigated whether their associations with cirrhosis are mediated through the underlying etiologies of CLD. Using two decades of longitudinal EHR data in MVP, we performed formal causal mediation analyses to estimate the direct effects of lead cirrhosis-associated variants on cirrhosis risk and the indirect effects operating through potential mediators, including alcohol use, Met-ALT, chronic viral hepatitis B or C, and other known causes of CLD. We selected cirrhosis-associated variants with suggestive evidence of association (P < 1.0×10^−5^) in the ancestry-specific discovery GWAS, namely 22 of 27 variants in the European and five in the African American GWAS (Supplementary Table 4).

We next evaluated these 22 variants in 11,738 European-ancestry MVP participants with cirrhosis and 444,882 controls, and five variants in 3,700 African American cirrhosis cases and 118,526 controls. A key prerequisite for mediation is that the exposure (here genetic variant) is associated with the mediator (e.g., CLD etiology); otherwise, there is nothing to mediate. We therefore first tested each variant for association with 9 major causes of CLD. In European Americans, 13 of 22 variants were associated with one mediator, whereas variants at five were associated with multiple mediators (P < 0.001), resulting in 26 variant-mediator pairs for formal mediation analysis (Supplementary Table 11). In African Americans, two of five variants were associated with a single mediator and three with multiple mediators at a nominal significance threshold (P < 0.05), resulting in nine variant-mediator pairs (Supplementary Table 12).

We next performed causal mediation analysis for cirrhosis on each variant-mediator pair using a counterfactual framework to estimate natural direct and indirect effects (Figure 3A, Methods). We observed near complete mediation (>70% of the variant’s effect on cirrhosis is explained by a mediator) at two loci in European Americans: ADH1B (87% mediated by alcohol-related disease) and *HFE* (73% mediated by hemochromatosis). Five additional variants in Europeans and three in African Americans showed evidence of partial mediation through a single CLD etiology (Figure 3A).

**Figure 3:**
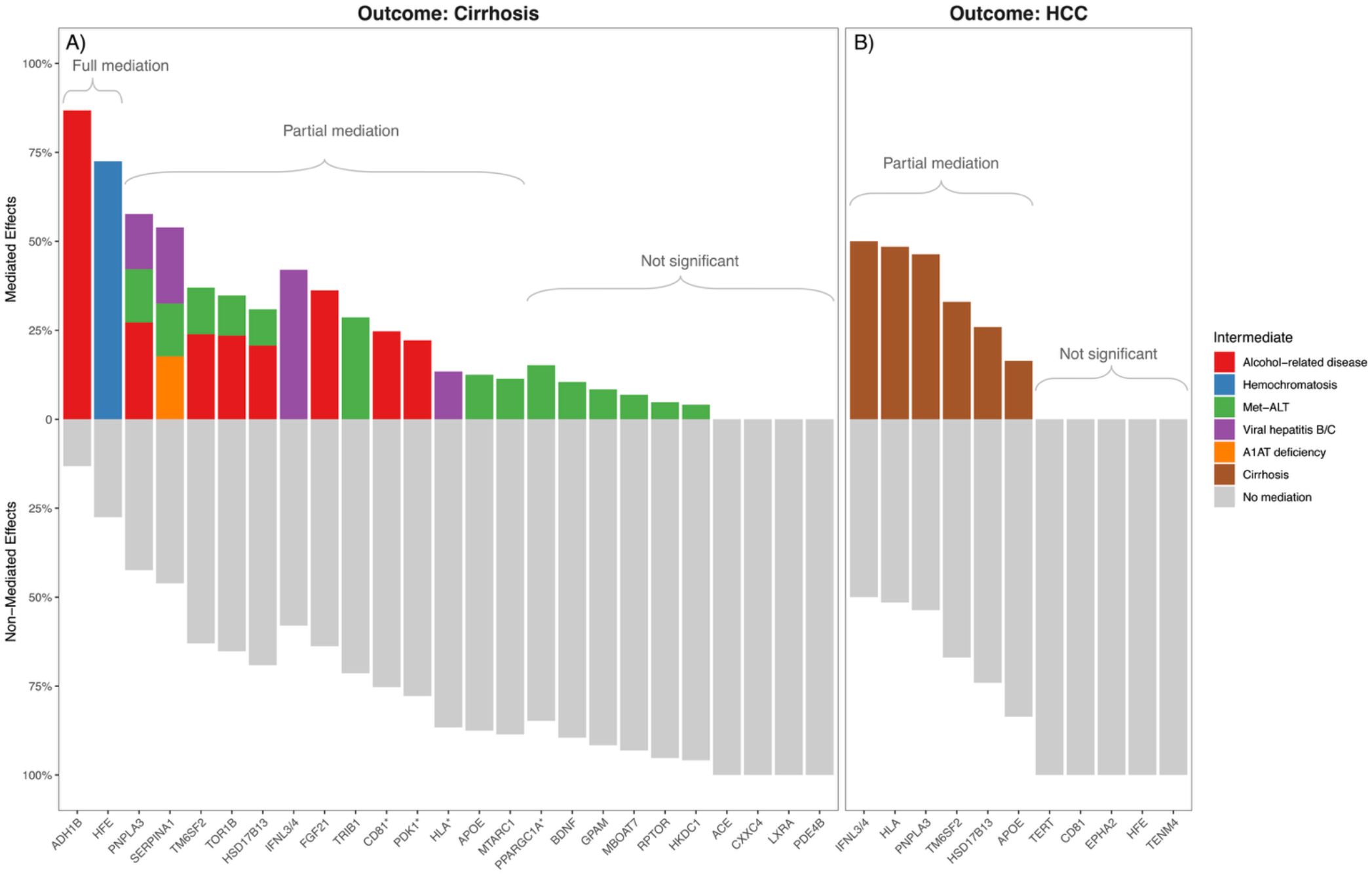
Mediation of genetic effects on cirrhosis and hepatocellular carcinoma. **(a)** Mediation analysis estimating the proportion of each SNP’s effect on cirrhosis that is mediated by specific liver disease etiologies. The top panel shows the percentage of the SNP effect mediated through indirect pathways, while the bottom panel displays the residual direct genetic effect on cirrhosis not explained by mediation. Percentage mediation is color-coded by etiology, namely alcohol use disorder, chronic viral hepatitis, metabolic dysfunction with elevated liver enzymes (Met-ALT), hemochromatosis, and alpha-1 antitrypsin deficiency (A1AT deficiency). Mediation results from African American analyses (*CD81, PDK1*, and *HLA*) are denoted with a star. **(b)** Mediation analysis of cirrhosis as a mediator between SNPs and HCC. The top panel shows the percentage of each SNP’s effect on HCC mediated through cirrhosis, while the bottom panel shows the direct effect on HCC independent of cirrhosis and other causes of liver disease. Namely, all models were additionally adjusted for covariates alcohol use, viral hepatitis, Met-ALT, hemochromatosis, alpha-1 antitrypsin deficiency, and other causes of CLD. Analyses were performed in European Americans.

Namely, the effect of *IFNL3/4* on cirrhosis was partially mediated by viral hepatitis, *FGF21* by alcohol-related liver disease, and *TRIB1, APOE, MTARC1* by Met-ALT (Supplementary Table 11). In African Americans, partial mediation was observed for *HLA* via viral hepatitis, and *CD81* and *PDK1* via alcohol use (Supplementary Table 12). Finally, five loci influenced cirrhosis risk through multiple pathways in European Americans (e.g., *PNPLA3, SERPINA1, TM6SF2, HSD17B13*, and *TOR1B*).

Collectively, these findings suggest that many genetic variants contribute to cirrhosis both directly and indirectly by increasing susceptibility to underlying causes of liver disease. This may explain why effect estimates appeared attenuated in the discovery GWAS using population controls. Notably, most SNPs retained residual direct effects on cirrhosis risk even after accounting for underlying etiologies, albeit with reduced magnitudes.

### Six loci partially impact HCC risk through cirrhosis

Because most HCC loci also confer risk for cirrhosis and cirrhosis is a key risk factor for HCC, we investigated whether their associations with HCC were mediated through cirrhosis. Using European-ancestry MVP participants (1,960 HCC cases and 457,737 controls), we performed mediation analysis treating cirrhosis as a mediator on the pathway from germline variation to HCC, while holding constant the effects of metabolic, viral, alcohol-related and other causes of CLD. Among European Americans in MVP, six of the 11 HCC variants were significantly associated with cirrhosis (P < 0.0001, e.g. *IFNL3/4, HLA, PNPLA3, TM6SF2, HSD17B13*, and *APOE*). All six variants showed evidence for partial mediation of HCC risk through cirrhosis but independent of an underlying etiology for cirrhosis (such as Met-ALT, alcohol use, or viral hepatitis) (Figure 3B, Supplementary Table 13). Supplementary Figure 3 provides an alternative visual representation of overlapping mediation pathways for cirrhosis and HCC.

### Genetic risk scores predict disease progression trajectories but not death

We next examined whether cumulative genetic risk from common variants influences the progression of liver disease through successive stages from CLD to cirrhosis and HCC. Using multi-state Markov survival models, we leveraged 20 years of longitudinal MVP data to model transitions between the following disease states: beginning at initial diagnosis of CLD (which could be Met-ALT, alcohol-related liver disease, chronic viral hepatitis, or another cause), progressing to cirrhosis, progressing to HCC, and ultimately to death. The assembled cohort included 134,841 European, 20,068 Hispanic, and 39,397 African American MVP participants with CLD at baseline. Supplementary Table 14 details the number of individuals who progressed to cirrhosis, developed HCC, or died with cirrhosis and/or HCC during the observation period within each ancestry.

We defined high genetic risk as having a cirrhosis GRS in the top 5^th^ percentile, based on 15 previously identified cirrhosis risk variants^15^, and compared outcomes to the remaining 95% of participants. Survival analysis demonstrated that a high GRS was associated with higher hazard ratios (HRs), indicating higher incidence rates and faster disease progression (Supplementary Table 15). In European Americans, a high GRS was associated with an almost two-fold increased HR of progressing from CLD to cirrhosis (HR 1.94, P = 2×10^−68^), and from cirrhosis to HCC (HR 1.65, P = 7×10^−8^). In Hispanic individuals, a high GRS also predicted progression from CLD to cirrhosis (HR 1.55, P = 4×10^−6^) and from cirrhosis to HCC (HR 1.46, P = 0.05). In African Americans, a high GRS was significantly associated with progression from CLD to cirrhosis (HR = 1.30, P = 6×10^−4^). Finally, a high GRS was not associated with mortality. Probability-of-state curves for cirrhosis and its underlying etiologies among European Americans are shown in Figure 4, with average times spent in each state provided in Supplementary Table 16.

**Figure 4:**
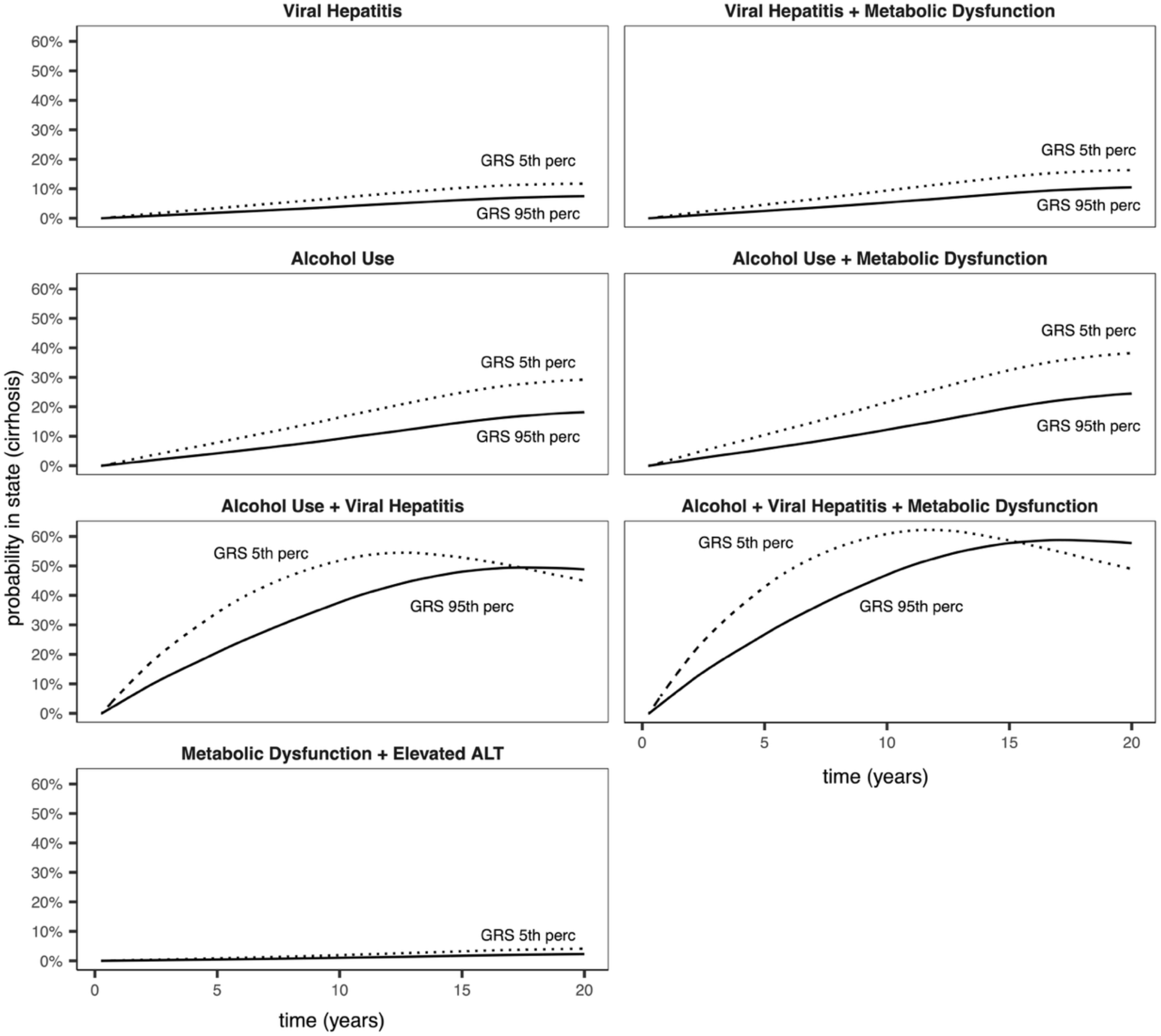
Probability of being in the cirrhosis state over time by etiology and genetic risk. Seven panels, stratified by combinations of etiologies (e.g. alcohol-related liver disease, chronic viral hepatitis, and metabolic dysfunction with elevated liver enzymes) show the probability of being in the cirrhosis state over a 20-year period following first CLD diagnosis. Probabilities were estimated using multi-state models with cirrhosis, HCC, and death as competing outcomes. Each panel compares individuals in the top 5th percentile (5^th^ perc) versus the bottom 95th percentile (95^th^ perc) of the genetic risk score. The x-axis shows time since first diagnosis of CLD in years, and the y-axis the probability of being in the state of cirrhosis at the respective timepoint. Across all etiologies, higher genetic risk was associated with an increased probability of cirrhosis. Among patients with both alcohol-related liver disease and viral hepatitis, a high genetic risk was further linked to earlier onset of cirrhosis.

**Figure 5:**
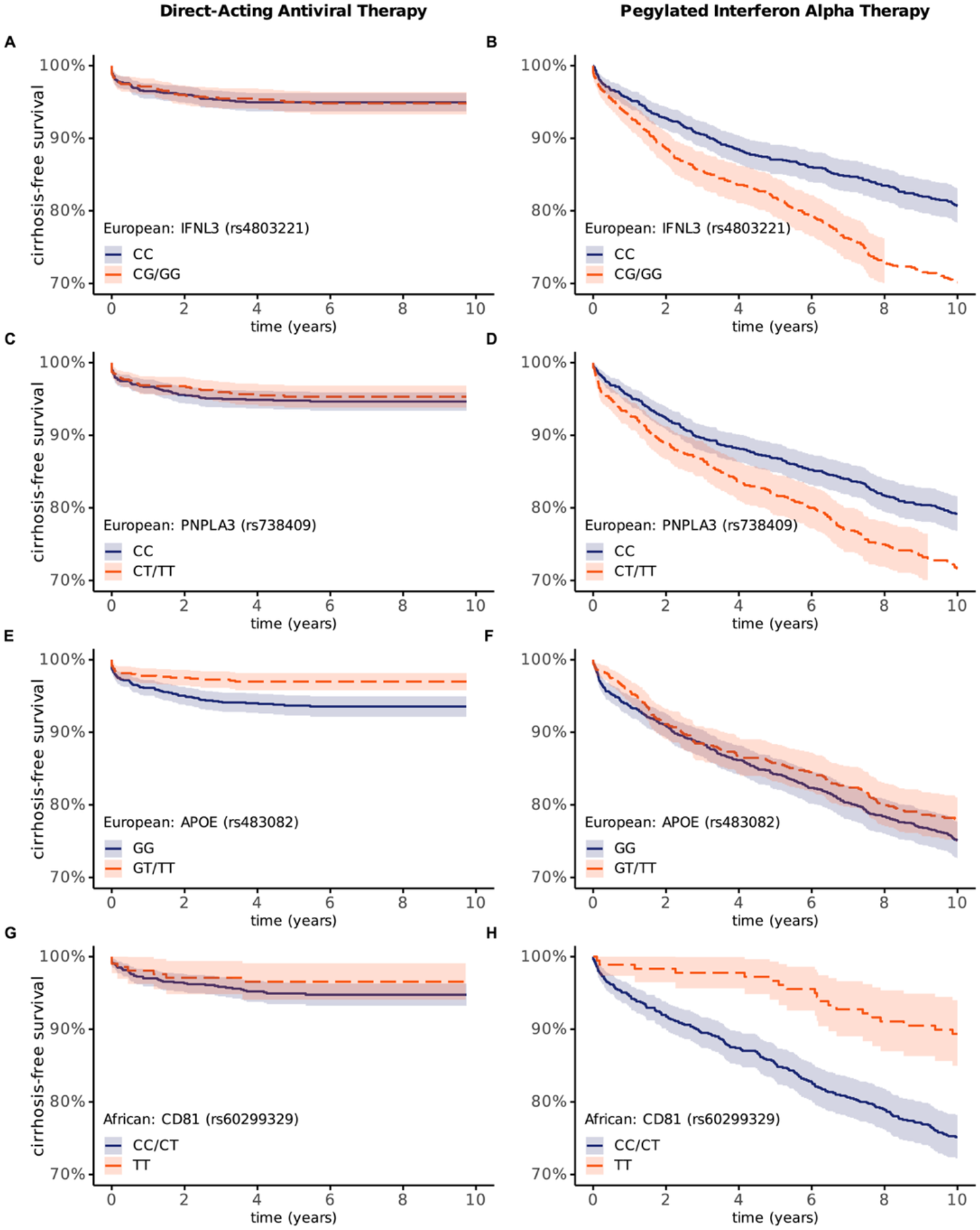
Cirrhosis-free survival by genotype and anti-viral treatment in chronic HCV infection. Eight panels display 10-year cirrhosis-free survival in treatment-naïve, non-cirrhotic individuals with chronic HCV infection, separated by ancestry and anti-viral treatment: direct-acting antivirals (A, C, E, G) and pegylated interferon alpha with ribavirin therapy (B, D, F, H). Kaplan–Meier curves compare difference in cirrhosis-free survival stratified by genotypes of IFNL3, PNPLA3, and APOE in European Americans, and CD81 in African Americans. The y-axis shows cirrhosis-free survival probability, and the x-axis time since treatment initiation in years.

Our analyses demonstrate that high genetic risk exerts a stronger effect on progression to cirrhosis than Met-ALT (HR 1.94 vs. 1.32, Supplementary Table 15). However, both are modest compared to effects of alcohol-related liver disease and viral hepatitis (HR of 12.6 and 5.4, respectively). By contrast, progression to HCC followed a different hierarchy of risk factors: viral hepatitis exerted the strongest effect (HR 6.4), followed by high genetic risk (HR 2.9), with alcohol contributing only modestly (HR 1.4). While viral hepatitis is a leading cause of cirrhosis worldwide with cirrhosis as a key risk factor for HCC, our findings show their differential role in driving cirrhosis and hepatocarcinogenesis.^7,27-32^

We further extended the multi-state framework to distinguish between compensated and decompensated cirrhosis as separate states. Decompensated cirrhosis is characterized by major complications largely related to portal hypertension, but also variceal hemorrhage, encephalopathy, and ascites as listed in Supplementary Table 2. Findings were largely concordant between analyses of overall cirrhosis and those distinguishing compensated from decompensated cirrhosis (Supplementary Table 17 and 18). Although high genetic risk for cirrhosis did not alter the likelihood of transition from compensated to decompensated cirrhosis, it was associated with higher hazards of progression from decompensated cirrhosis to HCC (P = 0.002) and to death (P = 0.034). Thus, while genetic risk for cirrhosis drives the development of cirrhosis and hepatocarcinogenesis, progression to cirrhosis decompensation mainly related to portal hypertension may be driven by other factors.

### Integrating clinical and genetic risk factors improves prediction of cirrhosis risk

We developed a multi-factorial prediction framework in MVP by integrating clinical risk factors with genetic risk scores. Specifically, we compared the predictive performance of a genetic risk score of total effects (GRS_total_), which capture overall polygenic variance, and a genetic risk score of direct effects (GRS_direct_), which are not mediated by intermediate phenotypes (see Methods). In total, seven logistic regression models were evaluated: Model 1 included only age and sex (baseline); Model 2 added clinical causes of CLD to the baseline model; Models 3 and 4 added GRS_total_ and GRS_direct_ to the baseline model, respectively; Models 5 and 6 added GRS_total_ and GRS_direct_ to the baseline + clinical model, respectively; and Model 7 further incorporated GRSs for indirect effects of underlying etiologies (e.g. GRS_alc_, GRS_met_, GRS_vhep_, GRS_aatd_, and GRS_hh_) into the baseline + clinical + GRS_direct_ framework.

A model including age, sex, and clinical covariates significantly outperformed the baseline model of age, and sex alone (AUC = 0.918 vs 0.628; Supplementary Table 19). When genetic risk scores were added the baseline model, both GRS_total_ (Model 3) and GRS_direct_ (Model 4) were significantly associated with cirrhosis (AUC = 0.643 and 0.642, respectively). The difference between the two was statistically significant, with the Akaike Information Criterion (AIC) favoring the model including GRS_total_ (P = 0.0001, Supplementary Table 20). These findings suggest that in the absence of clinical covariates, a GRS capturing total polygenic variance provides better fit than one restricted to direct genetic effects.

In contrast, when genetic risk scores were added models including clinical covariates (Models 5-6), GRS_direct_ showed a slightly higher AUC than GRS_total_ (0.921 vs 0.920; P = 0.0051), and a lower AIC, indicating improved fit. Finally, extending Model 6 with genetic scores for underlying etiologies (Model 7) led to a modest but statistically significant improvement (AUC = 0.922; P = 0.036). Together, these results demonstrate that GRS_total_ is more informative in models without clinical mediators, whereas combining GRS_direct_ and GRS_indirect_ enhances prediction in the presence of clinical risk factors.

### Proteome-wide drug discovery and colocalization identiXies drug targets for cirrhosis & HCC

We conducted a proteome-wide drug target Mendelian randomization (MR) analysis to identify circulating proteins that may causally influence cirrhosis or HCC risk, thus highlighting potential therapeutic relevance. This analysis leveraged summary statistics from a large proteomics GWAS that quantified plasma levels of ∼4,900 proteins in 35,559 Icelandic individuals. The MR screen identified multiple proteins whose genetically predicted levels were associated with disease risk (Supplementary Table 21). For cirrhosis, higher levels of PTPRJ and PARP1 were associated with reduced risk, suggesting potential for agonist-based therapeutic strategies. Conversely, elevated levels of IFNL3, ALDH2, and LCK were associated with increased cirrhosis risk, indicating that antagonist or inhibitory approaches may be warranted.

For HCC, higher levels of EPHA2, IGF2, GCKR, NCAN, and ITCH were associated with a reduced risk, supporting their potential as agonist-based targets, whereas increased NRBP1 expression was associated with higher risk, pointing to an antagonistic therapeutic strategy. Four targets were further supported by strong colocalization between protein levels and disease associations, namely PARP1, EPHA2, GCKR, and NRBP1. These findings provide a prioritized set of candidate proteins for validation to guide therapeutic development.

### Four variants are associated with differential cirrhosis risk after antiviral treatment

We compared host genotype-specific disease progression across two treatment eras of chronic HCV infection in MVP. The pegylated interferon (pegIFN)-based cohort included 1,859 European and 990 African American treatment-naïve, non-cirrhotic individuals treated with pegIFN-α and ribavirin between 2000 and 2010. Among them, 500 European and 242 African American patients developed cirrhosis within 10 years of treatment initiation (Figure 4, Methods).

The direct-acting antiviral (DAA) cohort comprised of 2,045 European and 1,083 African American treatment-naïve, non-cirrhotic individuals who initiated DAA therapy between 2013 and 2022, with 101 and 51 individuals, respectively, progressing to cirrhosis during the follow-up period (Figure 4, Methods).

In the pegIFN cohort, we observed pronounced genotype-specific differences on cirrhosis risk for *IFNL3/4, CD81*, and *PNPLA3* (Supplementary Table 22). Among European Americans, carriers of the *IFNL3/4* and *PNPLA3* risk alleles had poorer outcomes, reflecting interferon non-responsiveness associated with *IFNL3/4* and *PNPLA3*^33,34^. Furthermore, the minor allele of TOR1B was associated with improved cirrhosis-free survival. Among African Americans, we identified a novel association where carriers of the *CD81* risk allele exhibited reduced cirrhosis-free survival with pegIFN therapy.^35^ In a sensitivity analysis restricted to patients who received over 3 months of pegIFN/RBV therapy, the associations remained significant, and these variants were not associated with early treatment discontinuation (Supplementary Table 23).

In contrast, in the DAA-treated cohort, the effects of *IFNL3/4, PNPLA3*, and *CD81* on disease progression were attenuated, consistent with DAAs bypassing host genetic effects.^36^ We also observed a novel suggestive association in which carriers of the *APOE* minor allele demonstrated higher cirrhosis-free survival. Taken together, these findings underscore the role of host genetic variation in modulating differential risk of disease progression after interferon-based therapy and highlight the need to investigate potential genotype-dependent effects DAA treatment.

## DISCUSSION

Chronic liver disease is a growing global health burden, primarily due its progression to cirrhosis and HCC.^33,34^ Leveraging the largest multi-ancestry GWAS of cirrhosis and HCC to date, we identified 10 novel genetic associations for cirrhosis and three for HCC, providing new insights into the genetic architecture and pathogenic mechanisms of disease progression. These findings also carry potential therapeutic and prognostic implications.

### Control group selection and progression risk

Using population-based controls in disease progression studies risks misrepresenting the true *at-risk* population and may capture associations that reflect secondary effects of genetic predisposition rather than true progression drivers. Here, we systemically evaluated the impact of control group selection on genetic effect estimates. For cirrhosis, results were largely consistent between population and CLD controls. For HCC, however, using cirrhosis controls significantly attenuated associations at most loci. Equally important, most loci retained residual associations with progression in disease-restricted analyses, indicating that variants do contribute to progression, although precise effect size estimation will require modified study designs. Furthermore, we observed no reversal of effect direction between population and disease-restricted controls, reducing concern for collider bias, although this may not be generalized to all progression studies. Our findings emphasize the importance of careful control selection in studies of disease progression.

### Intermediate mechanisms of genetic risk

A key contribution of our study is the delineation of intermediate pathways through which genetic variants drive liver disease progression. Most cirrhosis-associated variants are established risk factors for underlying etiologies, such as alcohol-related liver disease, chronic viral hepatitis, metabolic dysfunction, hemochromatosis, or α-1-antitrypsin deficiency. For 15 cirrhosis loci, we formally demonstrated mediation through one or more of these etiologies, providing novel mechanistic insight. Similarly, six HCC loci exerted their effects on HCC through cirrhosis, independent of underlying etiology. Notably, three HCC-associated genes (e.g. *EPHA2, TERT*, and *TENM4*) that directly contribute to HCC risk are established cancer-related genes with roles in receptor tyrosine kinase signaling, telomerase regulation and senescence, and cellular differentiation, respectively^37-39^. At the same time, some loci (e.g. *HLA, IFNL3/4, PNPLA3, HSD17B13, TM6SF2, APOE*) contributed not only to underlying etiology for CLD but also to cirrhosis and HCC. Collectively, these loci may enhance risk stratification for liver disease progression.

### Integrated modeling of clinical and genetic risk

By combining genetic and clinical data, we developed an integrated risk model that improves cirrhosis prediction beyond the performance of either source alone. Importantly, our results highlight that the selection of how a GRS is constructed and weighted must be tailored to the specific analytic context: when used independently for population screening, a GRS should capture total genetic risk; whereas in models that already include clinical risk factors, it is preferable to exclude genetic effects mediated through those factors. Our comparison of models incorporating total, direct, and indirect genetic effects highlights this distinction: GRS_total_ performs better in models without clinical mediators, while GRS_direct_ and GRS_indirect_ together provide superior predictive accuracy when clinical covariates are included. This framework illustrates how partitioning genetic risk can optimize predictive modeling, enhance interpretability, and ensure appropriate application of GRS across different clinical and research contexts.

### Metabolic pathways and therapeutic implications

Several cirrhosis-associated genes converge on lipid-activated nuclear receptor pathways, including peroxisome proliferator-activated receptors and liver X receptors. We also identified fibroblast growth factor 21 (*FGF21*), PPARγ coactivator 1α (*PGC-1α*), and Liver X receptor alpha (*LXRα*) as novel cirrhosis-associated genes. *FGF21*, a fasting-induced hepatokine, activates PGC-1α^40,41^ to regulate glucose and lipid metabolism through downstream transcription factors (e.g., *PPARα, PPARγ, LXRα, FXR* and *HNF4α*)^42^. Phase II trials of FGF21 analogues have shown efficacy in resolving steatosis and improving fibrosis in patients with metabolic-associated steatohepatitis (MASH), including the SYMMETRY trial.^43-47^ Our mediation analysis indicates that *FGF21’s* effect on cirrhosis is mediated through alcohol consumption, consistent with reports of FGF21 analogues reducing alcohol intake in primates through liver-to-brain signaling axis involving neurons in the basolateral amygdala^48-53^. These findings suggest a potential role for FGF21 analogues in both metabolic and alcohol-related liver disease.

### novel virus-linked susceptibility loci: IFNL3/4 and CD81

We identified *IFNL3/4 (IL28B/IFNL4)* and *CD81* as novel susceptibility loci for both cirrhosis and HCC, which are involved in the hepatitis C virus (HCV) life cycle and disease pathogenesis. *IFNL3/4* polymorphisms influence HCV clearance during natural infection and on interferon-based antiviral therapy.^54,55,56^ *IFNL3/4* has also been linked to hepatic inflammation and fibrosis in non-viral CLD^57,58^. Our drug target MR nominated IFNL3 as a candidate drug target for cirrhosis. *CD81*, identified as a risk factor for cirrhosis and HCC in African Americans, encodes a tetraspanin critical for HCV cellular entry, cell-cell interactions, intracellular trafficking, immune signaling, and cancer progression.^59,60,61,62^ Our mediation analysis suggested that CD81 influences cirrhosis through alcohol-related pathways, consistent with animal models in which CD81 knockdown reduced alcohol-seeking behaviors^63^. Other mice models have shown that CD81 regulates adipose progenitor plasticity and beige fat formation, acting as a metabolic sensor that protects against obesity, insulin resistance, and adipose inflammation^64^.

### Protective effects of PNPLA3 and SERPINA1 against HCV infection

We observed that *PNPLA3* and *SERPINA1* variants were associated with reduced risk of HCV infection. Although initially counter-intuitive, these results are consistent with recent large-scale phenome-wide biobank meta-analyses (P < 0.05, https://platlas.cels.anl.gov/).^65^ In MVP, effect sizes were stronger, possibly reflecting a higher background HCV exposure as well as infection among U.S. Veterans.^66^ Mechanistically, PNPLA3-I148M may disrupt HCV replication by impairing lipid mobilization and altering lipid-droplet-Golgi dynamics^67-70^ needed for HCV lifecycle. For example, lower baseline viral load was reported in carriers of the 148M allele among individuals infected with HCV genotypes 1b and 2, but not genotype 3, which is independently associated with hepatic steatosis^71^. Similarly, *SERPINA1* mutations can impair the endoplasmic reticulum and Golgi apparatus^72^, thereby hindering HCV replication. In severe A1AT deficiency alleles, misfolded protein accumulates within the endoplasmic reticulum, inducing stress responses and reducing secretory efficiency. This convergence of impaired intracellular trafficking pathways may explain the observed protective effects of both *PNPLA3* and *SERPINA1* variants against HCV infection.

### Drug target discovery

Our MR-based drug target screen identified several proteins of therapeutic interest for cirrhosis including IFNL3. Furthermore, genetically increased EPHA2 activity was associated with reduced HCC risk, in contrast with the development of EPHA2 inhibitors as cancer therapeutics. This may reflect the dual rule of EPHA2 signaling: canonical pathways support tissue homeostasis and repair, whereas non-canonical signaling promotes tumorigenesis.^73,74^ These results highlight the importance of context-specific biology and suggest that EPHA2 activation, rather than inhibition, could be protective in pre-malignant liver disease, which is in line with our findings. The absence of an FGF21 signal for cirrhosis further emphasizes the importance of collecting disease-specific transcriptomic data in human tissues to enable the genetic evaluation of therapeutic mechanisms within their relevant biological setting.

### Genotype-dependent risk for liver disease progression to cirrhosis after antiviral treatment

Host genetics are known to influence the outcome of viral infection and treatment. Here, we show that *PNPLA3, IFNL3/4* and *CD81* variants differentially impact cirrhosis progression in HCV-infected patients who underwent PegIFN and ribavirin therapy, likely reflecting poor antiviral efficacy for PegIFN therapy which has been associated with these genetic variants. ^33,34 35^ For DAA therapy, however, such effects were markedly attenuated, likely because the very high cure rates with sustained viral clearance (>95%) minimize interindividual variability in treatment response.^75^ Nevertheless, carriers of the APOE minor allele showed improved cirrhosis-free survival following DAA treatment. The APOE ε4 allele has been previously associated with lower risk of HCV infection^76-78^, slower progression to advanced liver disease among HCV-infected individuals^79^, reduced fibrosis progression after liver transplantation^80,81^, and enhanced viral clearance and treatment response to pegIFN/ribavirin.^82^ In our study, the APOE minor allele was associated with a protective effect against cirrhosis in both DAA-treated and pegIFN-treated cohorts, with a stronger effect observed under DAA therapy. Our findings highlight the need to further investigate its role in the context of modern DAA therapy.

### Strengths and limitations

A unique strength of our study is the use of long-term longitudinal electronic health record data from MVP, which enabled detailed tracking of disease progression, estimation of mediating effects of underlying etiologies, and evaluation of treatment response, which is challenging in biobanks with predominantly cross-sectional data with high levels of missingness. Additional strengths include the use of disease-restricted controls to more accurately capture the at-risk population, the availability of high-quality immunogenetic imputation, and whole-genome sequencing data on over 100,000 participants, providing deeper resolution of genetic variation and enhancing discovery power. However, as with all EHR-based studies, potential misclassification of clinical diagnoses and residual confounding from unmeasured or imperfectly captured factors remain important limitations. These issues may influence effect size estimates, though the large sample size, replication across ancestries, and consistency of findings mitigate concerns about spurious associations.

## Conclusion

In sum, this study advances understanding of the genetic architecture and biological pathways underlying liver disease progression. We identify mechanisms by which variants influence progression, including through alcohol, metabolic, and viral pathways, highlighting potential therapeutic targets and demonstrating the value of integrating genetic and clinical data for precise risk prediction. While substantial work remains to translate these insights into targeted therapies and preventive strategies, our findings lay a strong foundation for future precision medicine approaches in liver disease.

## Data Availability

All data produced in the present work are contained in the manuscript, and corresponding genome-wide summary statistics are available through dbGAP (accession code phs001672.v12.p1) upon publication.

https://www.ncbi.nlm.nih.gov/projects/gap/cgi-bin/study.cgi?study_id=phs001672.v12.p1

---

**Supplementary Figure 1.**
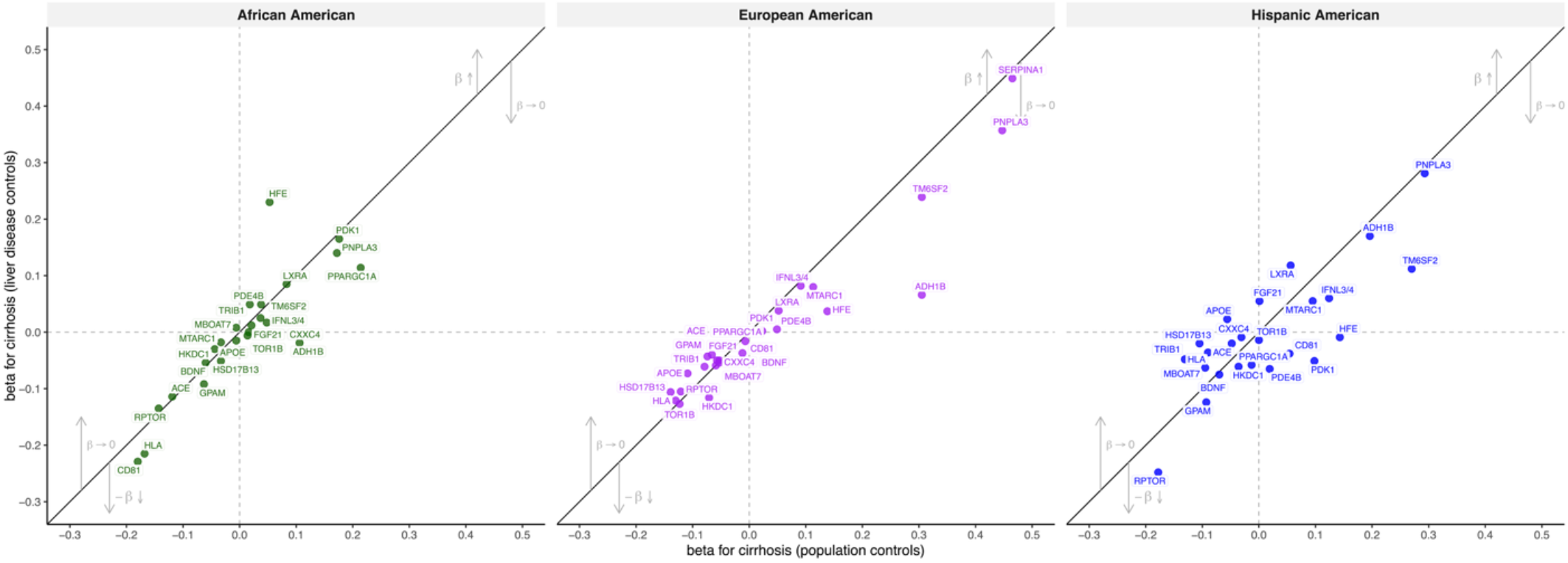
Scatterplots of SNP effect estimates for cirrhosis, stratified by ancestry (African American, European American, and Hispanic American). The x-axis shows the beta coefficient for cirrhosis estimated using population controls, while the y-axis shows the beta coefficient for cirrhosis estimated using chronic liver disease (CLD) controls. Each point represents a genetic variant (SNP).

**Supplementary Figure 2.**
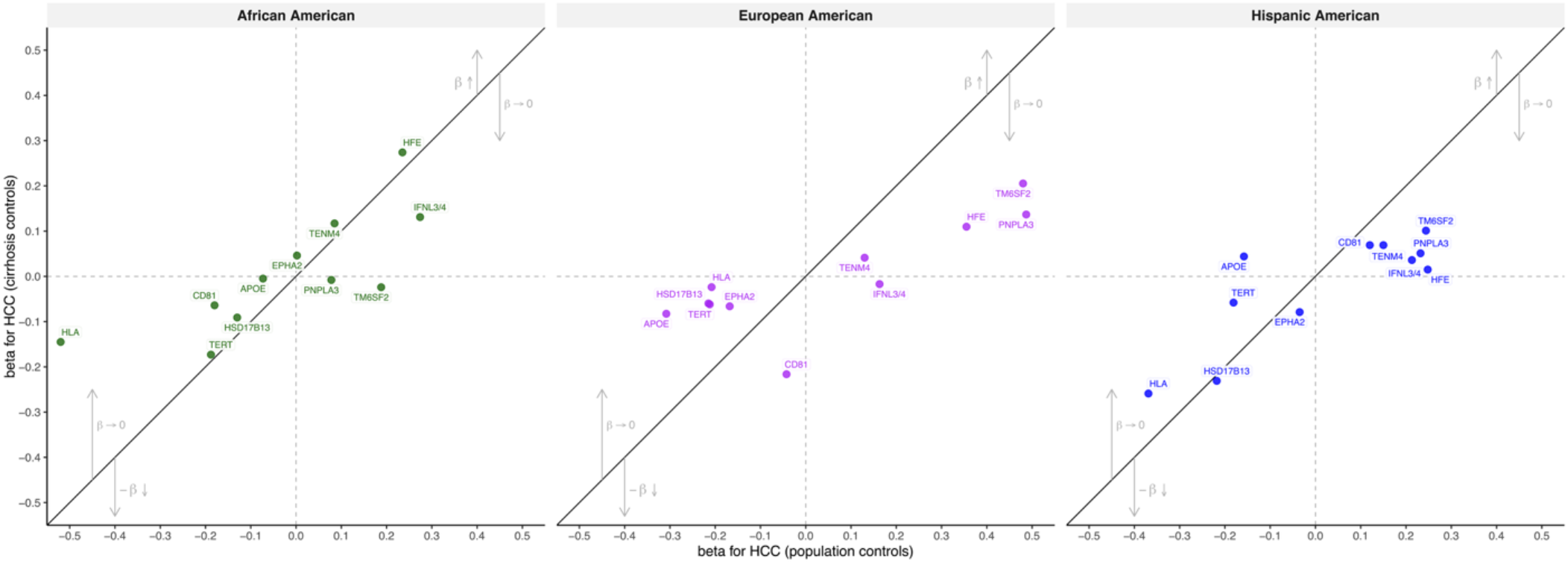
Scatterplots of SNP effect estimates for HCC, stratified by ancestry (African American, European American, and Hispanic American). The x-axis shows the beta coefficient estimated for HCC using population controls, while the y-axis shows the beta coefficient for HCC estimated using cirrhosis controls. Each point represents a genetic variant (SNP).

**Supplementary Figure 3.**
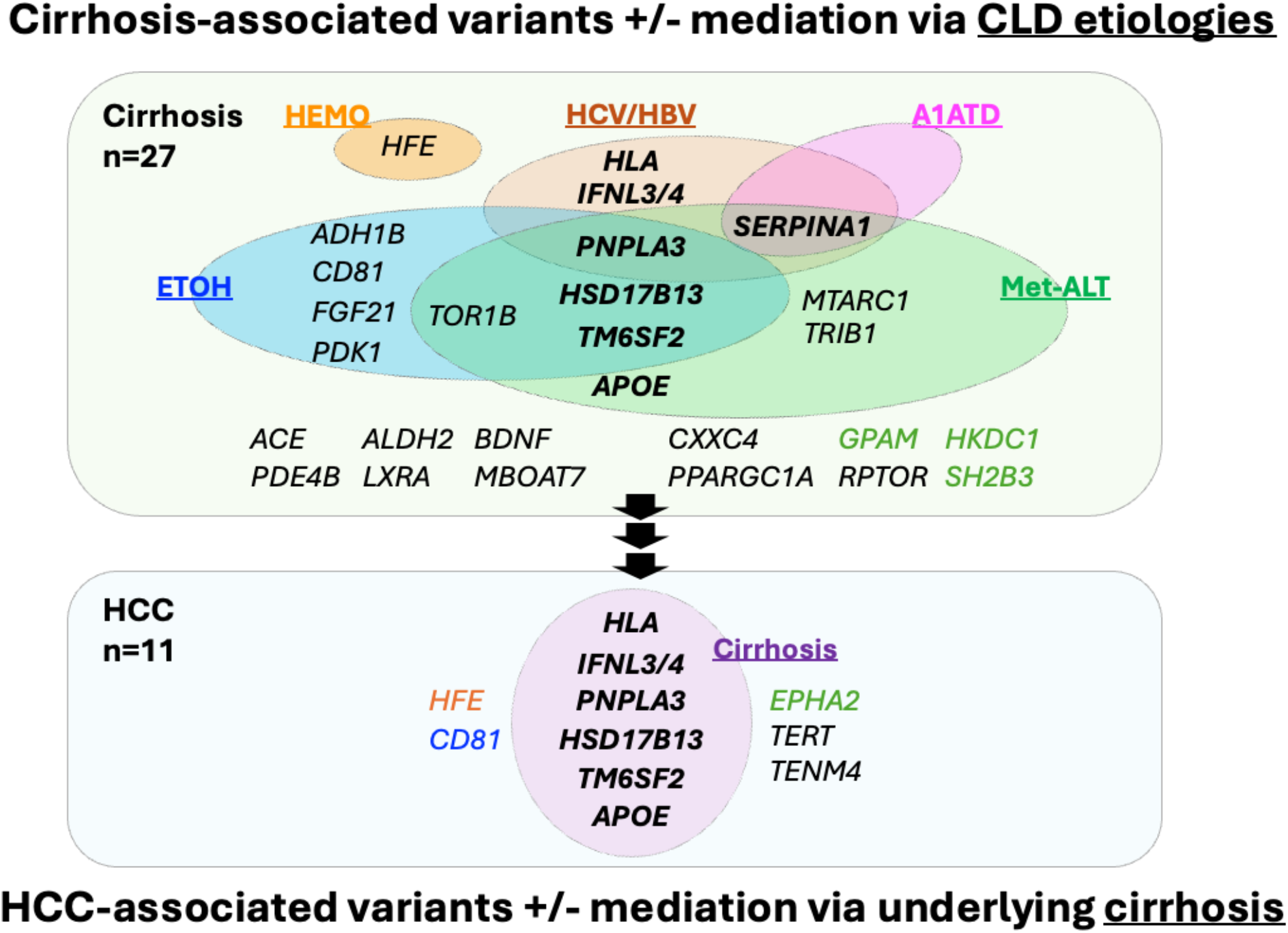
Twenty-seven SNPs were associated with cirrhosis, several of which mediate their effects through one or more underlying chronic liver disease (CLD) including alcohol-related liver disease (ETOH), hemochromatosis (HEMO), viral hepatitis (HCV/HBV), α1-antitrypsin deficiency (A1ATD), or metabolic-associated liver traits (Met-ALT). In addition, eleven SNPs were associated with hepatocellular carcinoma (HCC), some of which impact HCC risk indirectly by susceptibility to cirrhosis. *HLA, IFNL3/4, PNPLA3, HSD17B13, TM6SF2* and *APOE* show mediating effects for both cirrhosis and HCC.

## HUMAN RESEARCH PARTICIPANTS

### Ethics Oversight

All human research was approved within each contributing study by the relevant institutional review board and conducted according to the Declaration of Helsinki. MVP: VA Central Institutional Review Board (IRB). CHB-CID/DBDS: National Committee on Health Research Ethics; deCODE: National Bioethics Committee; Intermountain Healthcare: Intermountain Healthcare Institutional Review Board; UKB: Northwest Multicenter Research Ethics Committee; Geisinger DiscovEHR: The GHS project has received ethical approval from the Geisinger Health System Institutional Review Board under project no. 2006-0258; FinnGen: The Coordinating Ethics Committee of the Hospital District of Helsinki and Uusimaa; Estonian Biobank: Ethical approval 1.1-12/624 from the Estonian Committee on Bioethics and Human Research, Estonian Ministry of Social Affairs; Biobank Japan: Research ethics committees at the Institute of Medical Science, the University of Tokyo, the RIKEN Yokohama Institute, and the 12 cooperating hospitals; Copenhagen General Population Study and Copenhagen City Heart Study: institutional review boards and Danish ethical committees; All of Us: National Institute of Health All of Us Institutional Review Board; and Dallas Liver Cohort: University of Texas Southwestern Institutional Review Board. All participants (except for CHB-CID) provided written informed consent.

For CHB-CID, patients were informed about the opt-out possibility of having their biological specimens excluded from use in research in general. Since 2004, a national Register on Tissue Application (Vævsanvendelsesregistret) lists all individuals who have chosen to opt out and whose samples cannot be used for research purposes. Before initiating this study, individuals listed in the Register on Tissue Application were excluded.

## VA Million Veteran Program

We gratefully acknowledge the Veterans who participated in the Million Veteran Program as well as researchers and staff contributing to MVP recruitment, sample processing and data analyses. This research is based on data from the Million Veteran Program, Office of Research and Development, and Veterans Health Administration (MVP003/028) as well as MVP000. This publication does not represent the views of the Department of Veterans Affairs or the United States Government. This work was supported by grant number BX003362 (PI: K.M.C.) and used resources and facilities of the Department of Veterans Affairs (VA) Informatics and Computing Infrastructure (VINCI), including data analytics conducted by its Precision Medicine research team, which is funded under the research priority to Put VA Data to Work for Veterans (VA ORD 24-D4V-02). We thank Katie Pridgen, scientific writer, for reviewing the manuscript. D.E.K. is supported by I01-CX002010, M.V., J.P.J. and K.M.C. are supported by NIH/NIDDK R01-DK134575. B.F.V. is grateful for support for the work from the NIH/NIDDK (UM1-DK126194).

## Copenhagen Hospital Biobank Chronic Inflammatory Diseases and Danish Blood Donor Study (CHB-CID/DBDS)

We extend our sincere gratitude to the participants of the CHB-CID and DBDS for their invaluable contributions. We also thank the Department of Clinical Immunology at Copenhagen University Hospital for their support and collaboration. We acknowledge the dedicated staff of the biobank facilities and associated hospitals for their essential role in maintaining the infrastructure that made this study possible. This work was supported by BRIDGE—Translational Excellence Program (NNF20SA0064340 to J.G.), Beckett Fonden (23-2-10636 to J.G.), Independent Research Fund Denmark (Sapere Aude Research Leader, 9060-00012B to S.S.), Borregaard Clinical Ascending Investigator (NNF22OC0075038 to S.S.)

## UK Biobank

This research has been conducted using the UK Biobank Resource under Application Number 43247. We thank all UK Biobank participants and coordinators of the dataset. This work uses data provided by patients and collected by the NHS as part of their care and support. Full details of the study design and conduct are available on the UK Biobank website (https://www.ukbiobank.ac.uk). All participants gave their written consent and UK Biobank received ethical approval from the NHS Research Ethics Service (11/NW/0382).

## FinnGen

Data used in this research were obtained from FinnGen release 8, and we gratefully acknowledge the participants and investigators who contributed to the generation of this resource. The FinnGen project is funded by grants from Business Finland

(HUS 4685/31/2016 and UH 4386/31/2016), and by industry partners: AbbVie, AstraZeneca UK, Biogen, Bristol Myers Squibb (and Celgene Corporation & Celgene International II), Genentech, Merck Sharp & Dohme LLC, a subsidiary of Merck & Co., Inc., Rahway, NJ, USA, Pfizer, GlaxoSmithKline Intellectual Property Development, Sanofi US Services, Maze Therapeutics, Janssen Biotech, Novartis, and Boehringer Ingelheim. The following biobanks are acknowledged for delivering samples to FinnGen: Auria Biobank, THL Biobank, Helsinki Biobank, Biobank Borealis of Northern Finland, Finnish Clinical Biobank, Biobank of Eastern, Central Finland Biobank, Finnish Red Cross Blood Service Biobank, and Terveystalo Biobank. All Finnish biobanks are members of the BBMRI.fi infrastructure, and the FINBB is the coordinator of BBMRI-ERIC operations in Finland. The Finnish biobank data can be accessed through the Fingenious services managed by FINBB.

## Intermountain HerediGene

This research was supported by data and biospecimens from Intermountain Healthcare’s Precision Genomics Program. We gratefully acknowledge the participants, clinicians, and staff who contributed to the development and maintenance of the biobank and associated data resources. We also thank Intermountain Healthcare’s Precision Medicine team for their continued support of population-scale genomic research. Sequencing and genotyping for the initiative were performed in collaboration with deCODE genetics/Amgen.

## deCODE genetics

We used publicly available GWAS summary statistics provided by deCODE genetics (Reykjavík, Iceland), a subsidiary of Amgen Inc. We thank the participants in this study for their valuable contribution to research, and all investigators who contributed to data collection, phenotypic characterization of clinical samples, genotyping and analysis.

## Estonian Biobank

We thank all participants of the Estonian Biobank. The cohort is managed by the Institute of Genomics, University of Tartu, which is supported by the European Union through the European Regional Development Fund (project 2014-2020.4.01.15-0012 GENTRANSMED) and by the Estonian Research Council (grant PRG1911). Genotyping and data analysis were supported by the Estonian Genome Centre and its high-performance computing infrastructure.

## Biobank Japan

This study was conducted using data from the Biobank Japan Project. We thank all participants and the staff of the cooperating hospitals. The Biobank Japan Project was supported by the Ministry of Education, Culture, Sports, Science and Technology (MEXT) of Japan and the Japan Agency for Medical Research and Development (AMED). Summary statistics were obtained from the Biobank Japan PheWEB, which provides publicly available genome-wide association data. The Biobank Japan is supported by AMED under grant JP20km0605001.

## All Of Us Research Program

The All of Us is supported by the National Institutes of Health, Office of the Director—Regional Medical Centers (1 OT2 OD026549, 1 OT2 OD026554, 1 OT2 OD026557, 1 OT2 OD026556, 1 OT2 OD026550, 1 OT2 OD 026552, 1 OT2 OD026553, 1 OT2 OD026548, 1 OT2 OD026551 and 1 OT2 OD026555), IAA (AOD 16037), Federally Qualified Health Centers (HHSN 263201600085U), Data and Research Center (5 U2C OD023196), Biobank (1 U24 OD023121), The Participant Center (U24 OD023176), Participant Technology Systems Center (1 U24 OD023163), Communications and Engagement (3 OT2 OD023205 and 3 OT2 OD023206) and Community Partners (1 OT2 OD025277, 3 OT2 OD025315, 1 OT2 OD025337 and 1 OT2 OD025276). In addition, the All of Us Research Program would not be possible without the partnership of its participants. S.V. is supported by NIH R01 DK131033.

## Michigan Genomics Initiative

This research was conducted using data from the Michigan Genomics Initiative (MGI). We thank the participants of the MGI, as well as the faculty and staff of the University of Michigan Precision Health and Central Biorepository for their support and infrastructure. MGI is supported by the University of Michigan Medical School Central Biorepository, the University of Michigan Advanced Genomics Core, and Precision Health at the University of Michigan. Genotyping was performed at the University of Michigan Advanced Genomics Core. The MGI dataset includes electronic health record-linked genetic data made available through the University of Michigan Precision Health initiative.

## LITMUS Consortium/European SLD (MASLD) Registry^18^

This study was conducted using data and resources from the LITMUS project, which received funding from the Innovative Medicines Initiative 2 Joint Undertaking under grant agreement No 777377. This Joint Undertaking receives support from the European Union’s Horizon 2020 research and innovation program and EFPIA (European Federation of Pharmaceutical Industries and Associations). We thank all European SLD (MASLD) Registry partners and study participants for their valuable contributions to this collaborative effort. QMA is an NIHR Senior Investigator and is supported by the Newcastle NIHR Biomedical Research Centre, and the Horizon Europe, IMI2 and IHI research and innovation programs of the European Union under Grant Agreements 777377 (LITMUS), 101132901 (LIVERAIM), 101136259 (EDC-MASLD) and 101136622 (THRIVE). The views expressed are those of the authors and do not necessarily reflect the official position of IMI, the EU, or EFPIA.

The 17,781 population controls were recruited from the following sources: Wellcome Trust Case Control Consortium, (n=5159) typed on the Illumina Human1.2M-Duo; the Hypergenes cohort (http://www.hypergenes.eu/dissemination.html#pub), (n=1520) typed on the Illumina Human1M-Duo, KORA (https://epi.helmholtz-muenchen.de/), (n=1835) typed on the Illumina HumanOmni2.5Exome chip, and Understanding Societies (https://www.understandingsociety.ac.uk/), (n=9267) typed on the Illumina HumanCoreExome chip. Understanding Society: The UK Household Longitudinal Study is led by the Institute for Social and Economic Research at the University of Essex and funded by the Economic and Social Research Council. The survey was conducted by NatCen and the genome-wide scan data were analyzed and deposited by the Wellcome Trust Sanger Institute. Information on how to access the data can be found on the Understanding Society website https://www.understandingsociety.ac.uk/.

## Milano Biobank

We thank all participants and clinical staff involved in the Milano Biobank (Fondazione IRCCS Ca’ Granda Ospedale Maggiore Policlinico) for their invaluable contributions. The biobank is supported by the Italian Ministry of Health (Ministero della Salute), Ricerca Finalizzata 2021 RF-2021-12373889, Italian Ministry of Health, Ricerca Finalizzata PNRR 2022 “RATIONAL” PNRR-MAD-2022-12375656 (L.VC.V); Italian Ministry of Health (Ministero della Salute), Fondazione IRCCS Ca’ Granda Ospedale Maggiore Policlinico, Ricerca Corrente (DP); The European Union, H2020-ICT-2018-20/H2020-ICT-2020-2 program “Photonics” under grant agreement No. 101016726 - REVEAL (L.VC.V).

The European Union, HORIZON-MISS-2021-CANCER-02-03 program “Genial” under grant agreement “101096312” (L.VC.V, SP); Italian Ministry of Research (MUR) PNRR – M4 - C2 “National Center for Gene Therapy and Drugs based on RNA Technology” CN3, Spoke 4 “ASSET” (L.VC.V); PRIN 2022 MUR: “Disentangling genetic, epigenetic and hormonal regulation of Fe/heme metabolism in the gender-specific nature of NAFLD (DEFENDER)”.

## Penn Medicine Biobank

This research was conducted using data and biospecimens from the Penn Medicine Biobank (PMBB). We gratefully acknowledge the patients who consented to participate in the PMBB, as well as the faculty and staff who contributed to its operation. The PMBB is supported by the Perelman School of Medicine at the University of Pennsylvania and by the Penn Center for Precision Medicine. Genotyping and sequencing was performed with support from the Penn Center for Precision Medicine and Regeneron Genetics Center (Tarrytown, NY). This study also used linked electronic health record data maintained by Penn Medicine in support of the PMBB. We also thank the PMBB scientific advisory board and research community for ongoing support and collaboration. D.Y.Z. is supported by F30HL172382

## Regeneron Genetics Center

We thank the Regeneron Genetics Center (Tarrytown, NY) for generating the sequencing data used in this study. We also acknowledge the participants and the collaborating institutions that made this research possible. Whole-exome (or whole-genome) sequencing and initial data processing were performed at the Regeneron Genetics Center, Tarrytown, NY.

## BioVU Biobank

This research was conducted using the BioVU resource at Vanderbilt University Medical Center, which is supported by institutional funding and by the NIH-funded CTSA grant UL1 TR002243 from the National Center for Advancing Translational Sciences. We thank the participants whose de-identified DNA and clinical data made this research possible, as well as the Vanderbilt Institute for Clinical and Translational Research for infrastructure support. BioVU served as a replication resource for this study, and we acknowledge the contribution of the Vanderbilt research community to its maintenance and accessibility.

## Korea Biobank Project

This study was conducted using data from the Korean Genome and Epidemiology Study (KoGES), which is supported by the Korea Disease Control and Prevention Agency (KDCA). We thank all participants and staff members involved in the KoGES and the Korea Biobank Project. Data were obtained through the National Biobank of Korea (NBK), a part of the Korea National Institute of Health. Genotyping and quality control were conducted by the Korea Biobank Array Consortium. This study used publicly available genome-wide association summary statistics from the Korea Biobank Array Project, made available through the PheWeb portal by the KOGES. We thank the KDCA and the National Biobank of Korea for providing access to these data.

## Penn Transplant Cohort

Funding for this study was received from the Fred and Suzanne Biesecker Pediatric Liver Center at Children’s Hospital of Philadelphia (A.S.), Gift-of-Life Organ Procurement Organization (A.S.), and National Institutes of Health (NIH) National Institute of Allergy and Infectious Diseases (NIAID) grant no. U01AI152960-01 (A.S.).

## Geisinger DiscovEHR

This study was conducted in collaboration with the Geisinger Health System and the Regeneron Genetics Center. We thank all participants who provided samples. We thank the Geisinger-Regeneron DiscovEHR Collaboration team for their work in developing the data set used for analysis.

## Indiana Biobank

The biobank is a part of the Indiana Clinical and Translational Sciences Institute, a statewide collaboration of scientists at Indiana University, Purdue University, and the University of Notre Dame, as well as public and private partnerships. This study was funded, in part by Award Number UL1TR002529 from the National Institutes of Health, National Center for Advancing Translational Sciences, Clinical and Translational Sciences Award, and the National Center for Research Resources, Construction grant number RR020128 and the Lilly Endowment.

## European ALD-HCC Study

This work was supported by the Ligue Nationale contre le Cancer (Equipe Labellisée), an institutional grant from Bpifrance under the Hepatocellular Carcinoma Multi-technological consortium project, INSERM with the Cancer et Environnement (plan Cancer), Institut National du Cancer, the French Association for the Study of the Liver, Labex OncoImmunology (investissement d’avenir), grant IREB, Coup d’Elan de la Fondation Bettencourt-Shueller, the SIRIC CARPEM, FRM prix Rosen, Ligue Contre le Cancer Comité de Paris (prix René et André Duquesne), Fondation Mérieux Région Ile-deFrance, and Consortium HETCOLI (ITMO Cancer). The CiRCE study (ClinicalTrials.gov NCT01798173) was supported by the French National Research Agency under the programme Investissements d’Avenir (reference ANR-11-LABX-0021), Conseil Régional de Bourgogne, Fonds Européen de Développement Régional, Canceropole Grand-Est, Fondation de France. The SEPT9_CROSS study (ClinicalTrials.gov NCT03311152) was supported by the French Ministry of Health and Solidarity, Direction générale de l’offre de soins, French Eastern Interregional Group of Clinical Research and Innovation, University Hospital of Nancy/CRB lorrain (ClinicalTrials.gov NCT03311152), the Fonds Erasme for Medical Research, The Fonds Gaston Ithier for Oncology Research, a Collective Research Initiatives consolidator from the Université Libre de Bruxelles (EXPLORE project).

## German Cirrhosis Study

This study was supported by the German Ministry of Education and Research through the Virtual Liver Network, the PopGen 2.0 network biobank and institutional funds from the medical faculties of the Technical University Dresden and the Christian-Albrechts-University Kiel and the Swiss National Funds. The Community Medicine Research net of the University of Greifswald, Germany is funded by the Federal Ministry of Education and Research, the Ministry of Cultural Affairs and the Social Ministry of the Federal State of Mecklenburg-West Pomerania

## Japanese HCV-HCC study

This work was supported by a grant-in-aid from the Research Program on Hepatitis from the Japan Agency for Medical Research and Development (AMED JP21jk0210048, JP22jk0210103, JP25jk0210172) and the Ministry of Education, Culture, Sports, Science, and Technology (19H03640).

## METHODS

### Phenotype definitions of all-cause cirrhosis, HCC, and various causes of CLD

Cirrhosis cases and HCC cases were defined using ICD-9/10 diagnosis codes or cancer registry records in most cohorts. In MVP, we defined HCC using national cancer registry data and cirrhosis cases using a combination of ICD-9/10 codes and a laboratory-based fibrosis score, requiring Fib-4 > 2.67 at diagnosis to improve case specificity. Control definitions in MVP differed by analysis: for the primary discovery GWAS, controls were all individuals without a known history of cirrhosis (e.g. population controls), whereas in a secondary analysis, we used a restricted control population set to those at-risk for the respective outcome. For the cirrhosis GWAS, at-risk controls were individuals with CLD who had not developed cirrhosis, and for the HCC GWAS, at-risk controls were individuals with cirrhosis who had not developed HCC. A full description of each cohort’s case and control definitions is provided in Supplementary Tables 1 and 2.

### Multi-ancestry cohorts for common variant analysis of cirrhosis and HCC

We assembled association summary statistics from a total of 11 independent studies for cirrhosis and 12 studies for HCC, with MVP being the single largest contributor of cirrhosis and HCC cases. Genotyping in each study were assayed on various genotyping arrays, and standardized quality control at the sample and variant level was applied within each study (Supplementary Table 1). Within each study, participants were stratified by genetically inferred ancestry, and individuals identified as population outliers were excluded. Each study conducted an ancestry-specific GWAS for cirrhosis and/or HCC using logistic regression under an additive genetic model, adjusting for age, gender, and principal components of genetically-inferred ancestry using PLINK, REGENIE, or SAIGE software^83^.

Genotype imputation in each study was performed using one of several reference panels (NHLBI TOPMed reference panel, 1000 Genomes Project, or Haplotype Reference Consortium^84-86^). After imputation, variants were filtered using standard quality control thresholds: Hardy-Weinberg equilibrium P > 1 × 10^−10^, INFO imputation accuracy score > 0.3, genotyping call rate >0.975, and minor allele frequency >1% in the relevant ancestry group. The effect size estimates (log-odds ratios) from the regression analysis represent the log-odds change in disease risk per copy of the effect allele while holding other independents (e.g., covariates) in the model constant.

### Multi-ancestry meta-analysis of common variants for cirrhosis and HCC

We combined the ancestry-specific association results through inverse-variance weighted fixed-effects meta-analysis of log-odds ratios as implemented in METAL^87^. Variants were considered genome-wide significant if they passed the conventional P-value threshold of 5×10^−8^. We used an LD- and distance-based clumping procedure (500 kb window and/or LD *r*^2^ >0.05 in the relevant ancestry using 1000 Genomes as a reference panel) to defined approximately independent loci from the meta-analysis results. Multi-ancestry and ancestry-specific summary statistics are displayed in Table 1 and 2, Supplementary Tables 4 and 6, and their corresponding genome-wide summary statistics are available through dbGAP (accession code phs001672.v12.p1).

To assess potential inflation of effect estimates when using population controls, we performed disease-restricted analyses within the Million Veteran Program (MVP). For cirrhosis, controls were restricted to individuals with CLD but without cirrhosis. For HCC, controls were restricted to individuals with cirrhosis but without HCC. We then compared effect estimates from the primary GWAS with population controls to those obtained in the disease-restricted analyses for the lead ancestry-specific loci. Differences in effect estimates between population and disease-restricted controls were quantified using z-scores, and statistical significance was assessed by calculating corresponding P-values which are listed in Supplementary Tables 4 and 6.

### Replication of common variants associated with cirrhosis and HCC

To validate the genome-wide association study (GWAS) findings, we performed an independent replication analysis for cirrhosis and HCC with population controls using data from multiple external cohorts. We assembled a meta-analysis dataset comprising several independent studies (Supplementary Tables 5 and 7), each with genotyped or imputed genetic data, to evaluate the associations of the lead variants identified in the discovery phase. Association testing was performed first within each replication cohort using logistic regression under an additive genetic model, adjusting for age, sex, and ancestry principal components. We then meta-analyzed the results across all replication cohorts using inverse-variance weighted fixed-effects meta-analysis, as implemented in METAL. We assessed replication significance using a Bonferroni-corrected threshold (P<0.0018 for cirrhosis and P<0.0045 for HCC), with secondary analyses examining directional consistency and nominal significance (P <0.05). Results from the common variant replication analysis are shown in Table 1.

### Immunogenetic discovery analysis for cirrhosis and HCC using HLA predicted genotypes

Given HLA critical role in immune-mediated liver injury and cancer surveillance^88-90^, we performed a detailed analysis of imputed HLA genotypes with cirrhosis and HCC in MVP. Using the HIBAG algorithm, we imputed four-digit HLA alleles for three class I genes (e.g., HLA-A, HLA-B, and HLA-C) and four class II genes (e.g., HLA-DPB1, HLA-DQA1, HLA-DQB1, and HLA-DRB1) from hard-called MVP genotype data^91,92^. We applied pre-trained HIBAG models based on the Affymetrix Axiom UK Biobank Array, which is well-matched to the MVP genotyping array which covers more than 95% of the training variants. Ancestry-specific imputation parameters were used: European-ancestry MVP samples were imputed with European reference models, and African American and Hispanic MVP samples with a multi-ethnic model (HLARES + HapMap). We retained common HLA alleles (those with minor allele count >50 within each ancestry) for association analysis, resulting in 203 HLA genotypes.

We tested each HLA genotype for association with cirrhosis and HCC in each ancestry using logistic regression (additive allele dosage model). Covariates included age, gender, the first 10 principal components of genetic ancestry, and indicator variables for the presence of various CLD etiologies (Met-ALT, alcohol use disorder, chronic viral hepatitis B/C, alpha-1 antitrypsin deficiency, hemochromatosis, autoimmune hepatitis, Wilson’s disease, primary biliary cholangitis, and primary sclerosing cholangitis). This approach accounts for potential confounding by these factors on the HLA-disease associations. We report all associations in the HLA-region for cirrhosis and HCC in Supplementary Table 9.

### Variant calling from WGS data in the Million Veteran Program

The Million Veteran Program (MVP) has generated deep whole-genome sequencing (WGS) data for 109,826 participants. We processed the raw WGS data by adapting the Broad Institutes “$5 genome” GATK pipeline (https://github.com/gatk-workjlows/five-dollar-genome-analysis-pipeline)^93^. Briefly, sequence reads were aligned to the GRCh38 human reference genome for mapping and variant calling using BWA-MEM (v0.7.15), and alignments were then compressed to CRAM and stored in a lossless manner^94^. Variant calling was performed with GATK 4.1.0.0 in gVCF model, which produces a per-sample GVCF containing all reference call blocks, single nucleotide variants (SNVs), and short insertion and deletions (INDELs)^95^.

We implemented comprehensive quality control (QC) at multiple levels. Sequencing-level QC consisted of FastQC (v0.11.4) to assess base sequence quality, read length, and GC content. Alignment-level QC included the use of SAMTools flagstat (v0.1.19) was used to quantify mapping statistics (e.g. number and percentage of properly mapped reads and properly paired reads)^96^ as well as verifybamID (GATK) to estimate DNA contamination rates for individual genomes^95^. Variant-level QC included the use of RTG Tools (vcfstats) to compute per-sample variant counts, transition/transversion ratios, and heterozygosity ratios^97^

Building on best practices from large biobank sequencing efforts, we implemented stringent filters at the variant, sample, and genotype level. For sample-level QC, we required: per-base sequence quality score ≥28, GC content within 40%-41%, ≥95% properly mapped reads, and estimated contamination <5%. We also required a sample call rate ≥97% and mean sequencing depth ≥18× for a sample to be included. For genotype-level QC, we set thresholds depending on genotype type: reference homozygous alleles required a genotype quality ≥20 (Phred scale); alternate homozygous alleles required ≥90% of reads supporting the alternate allele, genotype quality ≥20, a reference allele frequency ≤50%, and a reference allele likelihood ratio (Phred) ≥ 20; heterozygotes required a reference allele likelihood ratio ≥20, and an alternate allele fraction ≥20% (with alternate and reference allele depths summing to ≥90% of total depth). Finally, at the variant level, we required that each variant had at least one alternate allele in the dataset and an overall variant call rate ≥80%. After QC, 102,677 individuals and 663,351,127 variant calls remained for analysis.

### Annotation of sequencing-based variant calls

We annotated all 663,351,127 variants from WGS using Ensembl Variant Effect Predictor (VEP v110^98^), focusing on functional consequences in protein-coding genes. We were particularly interested in rare, high-impact variants. Predicted loss-of-function (pLoF) variants were defined as variants expected to severely disrupt gene function and include (a) insertions or deletions resulting in a frameshift, (b) insertions, deletions or single nucleotide variants resulting in the introduction of a premature stop codon or in the loss of the transcription start site or stop site, and (c) variants in donor or acceptor splice sites. We restricted pLOFs to canonical transcripts in protein coding genes and further required that LOFTEE (Loss-Of-Function Transcript Effect Estimator) flagged them as “High Confidence” pLOFs^99^. Rare deleterious missense variants were defined as missense mutations predicted to be “likely pathogenic” by AlphaMissense^100^, again restricting to canonical transcripts. We used a 1% minor allele frequency threshold to define “rare” for both pLoFs and deleterious missense variants.

### Gene-burden discovery analysis of rare deleterious variants for cirrhosis and HCC

We performed gene-burden association tests to identify genes in which rare variants contribute to cirrhosis or HCC risk. Separate tests were done within each ancestry group (European and African American) using the SAIGE-GENE+^83^ framework^101^. We applied the SKAT, SKAT-O, and Burden test with small-sample adjustments and a hybrid Firth correction as needed, using the “minimum P-value” method to combine results across variant sets for each gene. For each gene, we defined two sets of qualifying variants based on the annotations described above and minor allele frequency in that ancestry: 1) high-confidence pLoF variants with MAF < 1% and 2) high-confidence pLoF plus likely-pathogenic missense with MAF < 1%. We included age, gender, and first 5 principal components of genetic ancestry as covariates in each gene-level test. All significant gene-level associations from the discovery analysis are shown in Supplementary Table 10.

### Replication cohorts for coding variant and gene-burden analyses for cirrhosis and HCC

To validate the gene-burden associations identified in the discovery phase, we conducted independent replication analyses using whole-genome sequencing (WGS) and exome-sequencing data from multiple external cohorts. The replication datasets included the UK Biobank (UKBB), Mass General Brigham Biobank (MGB), All of Us Research Program, Milano Biobank, and the Regeneron Genetics Center (RGC). Gene-burden association analyses were conducted separately within each replication cohort using GENESIS, SAIGE-GENE+ or REGENIE. Association tests were performed under an additive genetic model, adjusting for age, sex, and first 5 genetic principal components. Replication results were meta-analyzed across cohorts using inverse-variance weighted (IVW) fixed-effects meta-analysis, implemented in METAL. Significance thresholds were set using Bonferroni correction to account for multiple testing, and P <0.05 considered nominally significant. Directional consistency between discovery and replication cohorts was also assessed using a binomial test. The results of the gene-burden replication analysis are shown in Supplementary Table 10.

To validate the coding variants identified in our GWAS, we leveraged whole-exome sequencing data from the Regeneron Genetics Center (Supplementary Table 8). The RGC dataset provides high-depth sequencing data across multiple ancestries, allowing for independent replication of coding variants associated with cirrhosis and hepatocellular carcinoma (HCC). Association tests were performed separately within each ancestry using REGENIE, adjusting for age, sex, principal components and relatedness and the meta-analyzed using fixed-effect inverse variance weighted effects in METAL.

### Causal mediation analysis of underlying etiologies between genotype and outcomes

We applied causal mediation analysis to quantify the extent to which genetic variants influence cirrhosis risk indirectly via intermediate phenotypes (here, alcohol-related liver disease, metabolic dysfunction with elevated liver enzymes, viral hepatitis, alpha-1 antitrypsin deficiency, hemochromatosis, auto-immune hepatitis, Wilson’s disease, primary biliary cholangitis, and primary sclerosing cholangitis). We selected 22 variants in European Americans and 5 variants in African Americans that showed a marginal association with cirrhosis and HCC at P<1.0×10^−4^ (see Supplementary Table 4 and 6). To test for variant–mediator associations, we first ran logistic regressions of these variants on each cause of CLD, with covariates of age, sex, first 5 principal components of genetic ancestry. We moved forward with variant–mediator pairs that met significance criteria (P<0.001 in European Americans, P <0.05 in African Americans) for formal counterfactual mediation analysis with cirrhosis as the outcome (Supplementary Table 11 and 12).

For each qualifying variant–mediator pair, we used the medflex package in R to fit natural effects models for nested counterfactuals for cirrhosis risk using the imputation approach method^102-104^. This method decomposes the total effect of a variant on cirrhosis into a natural direct effect (not through the mediator) and a natural indirect effect (through the mediator). Indirect effects denote the part of the total effect of SNP that is mediated by a cause of liver disease, while direct effects reflect the rest of the total effect. We specified a logistic regression for the cirrhosis outcome, including the variant, the mediator, and confounders of age, gender, PCs and other causes of liver disease. We obtained estimates and robust standard errors for the natural direct and indirect effects based on the sandwich estimator. We defined partial mediation when the indirect effect is statistically different from the total effect using the Z-score statistic and associated two-sided P-value, and full mediation if >70% of the observed effect between variant and cirrhosis is mediated through an intermediate (Figure 3, Supplementary Tables 11 and 12).

For HCC, we conducted counterfactual-based mediation analysis to quantify the extent to which the effect of each genetic variant is mediated through cirrhosis. In this framework, the variant served as the exposure, cirrhosis as the mediator, and HCC as the outcome. All 11 HCC loci were evaluated. Covariates included age, sex, the first ten principal components of genetic ancestry, and clinical factors associated with liver disease: alcohol-related liver disease, viral hepatitis, metabolic dysfunction with elevated liver enzymes, and hemochromatosis. We estimated natural indirect effects, natural direct effects, and the proportion mediated. Standard errors and confidence intervals were derived using robust sandwich estimators as implemented in the R medflex package. Results are shown in Figure 3 and Supplementary Table 13.

### Construction of a genetic risk score that captures total polygenic variance for cirrhosis

To construct a genetic risk score (GRS) for cirrhosis, we extracted the lead variants and accompanying effect size weights from a recent independent large cirrhosis GWAS by Ghouse *et al*.^15^ The lead 15 lead variants for cirrhosis in that study are located in or near PDE4B, MTARC1, ZFP36L2/HAAO, GYPC, HSD17B13, ADH1B, HFE, HLA, TRIB1, GPAM, ALDH2, SERPINA1, TM6SF2, APOE, and PNPLA3 and were all available in MVP. We selected the 15-variant GRS as it outperformed their expanded 36-variant GRS from an endophenotype-based analysis. For each MVP participant, we calculated the GRS as the sum of the risk allele count at each locus weighted by the published log-odds ratio (Supplementary Table 15 and 18).

### Multi-state survival analysis of genetic risk scores on cirrhosis, HCC, and death

We used multi-state survival modeling to evaluate the impact of cumulative genetic risk on progression through clinical disease stages. We defined four states: (1) CLD without cirrhosis or HCC, (2) cirrhosis, (3) HCC, (4) death. Transitions between these states included: CLD → cirrhosis, CLD → HCC, CLD → death, cirrhosis → HCC, cirrhosis → death, and HCC → death. Individuals may either progress to the endpoint through several intermediate stages or directly move to death, where we treated death as an absorbing state (no transitions out once reached). If an individual with CLD dies with no intermediate events of cirrhosis or HCC (CLD → death), then such a death is considered to have occurred due to causes not related to liver diseases; otherwise, it is considered all cause-mortality after cirrhosis or HCC. We used a semi-Markov approach which allows transition rates to depend on time since entry into the current state, as implemented in the mstate package in R.

The timepoint of study entry, or time zero, for each individual corresponded with the date of first diagnosis of chronic liver disease of any cause within the health record from January 1^st^ of 1999 onwards (metabolic dysfunction with elevated liver enzymes, alcohol-related liver disease, viral hepatitis B or C, and cases of other causes of liver disease). Individuals were censored at date of death or last follow-up (December 31^st^, 2021), or at 20 years of follow-up, whichever came first.

We estimated 20-year risks using cause-specific hazard function to analyze the direct association of GRS with the instantaneous hazard of the respective events. We fitted transition-specific Cox proportional hazards models for each of the six possible transitions, allowing each transition to have its own baseline hazard function. GRS was included as an independent variable and dichotomized into the top 5^th^ percentile (highest genetic risk) and bottom 95^th^ percentile. Baseline covariates included age at CLD diagnosis, sex, and indicator variables for the presence of alcohol-related liver disease, metabolic dysfunction with elevated liver enzymes, chronic viral hepatitis B or C, and other and other less common causes of CLD (which were grouped together for the analysis). We allowed for each transition to have a separate baseline hazard and that the effects of the covariates could differ for each transition. All analyses were performed using R (version 4.0) using the coxph function and the mstate package^105^ for handling multi-state data. Transition-specific results for the GRS and all covariates (hazard ratios, confidence intervals, P-values) are shown in Supplementary Table 15.

To estimate the state occupation probabilities and expected time spent in each state over 20 years, we used the Aalen-Johnson estimator and calculated these quantities for several frequent prototypical clinical profiles, namely: (1) Met-ALT only (no alcohol or viral hepatitis), (2) viral hepatitis only (no alcohol or Met-ALT), (3) viral hepatitis + Met-ALT (no alcohol), (4) alcohol (no viral hepatitis or Met-ALT), (5) alcohol + Met-ALT (no viral hepatitis), (6) alcohol + viral hepatitis (no Met-ALT), and (7) alcohol + Met-ALT + viral hepatitis. For each of these profiles, other causes of liver disease were assumed absent. These profiles correspond to combinations of the etiological covariates in our model. We then calculated the probability of each state (CLD, cirrhosis, HCC, death) at 20 years and the mean duration spent in each state. These results are presented in Supplementary Table 16.

We also extended the model to include a fifth state for decompensated cirrhosis (defined by complications such as variceal bleeding, hepatic encephalopathy, or ascites). This introduced additional transitions (e.g., cirrhosis → decompensated cirrhosis, and decompensated cirrhosis → HCC). We refit the models accordingly (Supplementary Tables 17 and 18). The effects of the GRS were examined in this expanded model to see if genetic risk influenced transitions to or from decompensated cirrhosis.

### Construction of genetic risk scores of direct and indirect genetic effects

To further explore and utilize the distinction between direct and mediated genetic effects, we constructed separate GRS for direct and indirect effect alleles to allow for genetic variants to exert their risk of disease progression via underlying mediating risk factors. Consider we start with a classic regression model without mediators, where the outcome of interest is binary to fit the observed data:

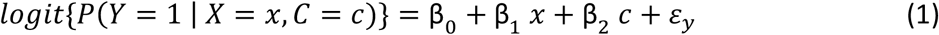

where *Y* is the vector of a binary trait for *N* individuals; X is the vector of genotypes; and C is a vector of covariates (such as sex, age, and global ancestry proportions). We assume a fixed effects regression model, where B is the vector of additive effects of the independent variables including an intercept, and e is the residual effect. Using this as a training model, then the genetic risk score for explaining total polygenic variance for risk of disease is defined as the sum of weighted alleles:

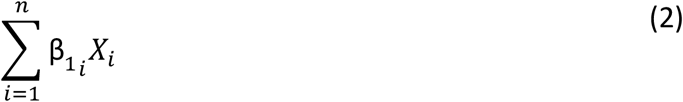

Here, it is assumed that the genetic liability of a disease is quantified by an additive model that combines the risk alleles into a single score (e.g., construct validity). When the variants are used to predict disease in the absence of non-genetic risk factors, then the genetic score rightly needs to capture total genetic variance. However, if we are interested in including genetic risk in a prediction model with clinical risk factors that mediate part of the polygenic risk, then the genetic score needs to capture the polygenic risk that is not captured by the intermediate phenotypes (e.g. direct genetic effects rather than total genetic effects). The content validity of the sum of weighted alleles by total effects is therefore valid in a model without clinical covariates but not in a prediction model that includes mediating clinical risk factors between genotype and disease. In the case of liver cirrhosis, we showed with mediation analysis that several genetic variants partially or fully exert their effect on cirrhosis via intermediate clinical risk factors, such as alcohol-related liver disease, metabolic dysfunction with elevated liver enzymes, and chronic viral hepatitis.

Therefore, to construct a genetic risk score of direct effects, we will conceptualize a regression for disease that includes both the genotype as well as the mediator as independents and eliminate the effect that the independent has on the outcome via the mediator.

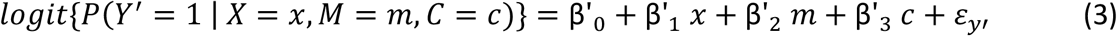

where *Y* is the vector of a binary trait for *N* individuals; X is the vector of genotypes; M is the vector of a mediator, and C is a vector of covariates. And now B represent the direct genetic effect of the variant on outcome (while the M is held constant), which can serve as weights for a GRS that captures direct effects only:

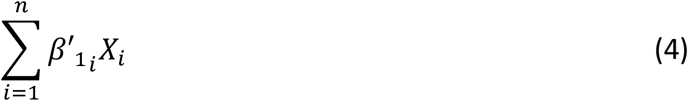

The direct effect GRS should be used in a prediction model where the mediator is included as a predictor. We can also expand on this genetic model and additionally include the indirect effects separately which is ultimately a weighted sum of direct effects on the mediator multiplied by the value of the mediator. First, we model the mediator as a function of the genetics:

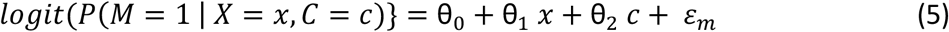

where *M* is the vector of a binary mediator for *N* individuals; X is the vector of genotypes; and C is a vector of covariates. The indirect effect of SNP on outcome via the mediator is defined as the product of the effect of the variant on the mediator () times the effect of the mediator on the outcome (), as follows:

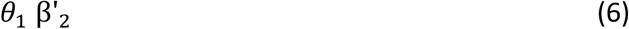

The risk model integrating clinical covariates, direct effects, and indirect effect of a single mediator is defined as follows:

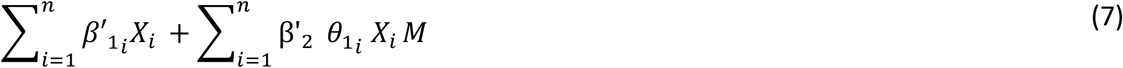

Because the sum of a constant times a variable equals to the constant times the sum of the variable, and the sum of a constant is equal to times the constant, the formula can be rewritten as:

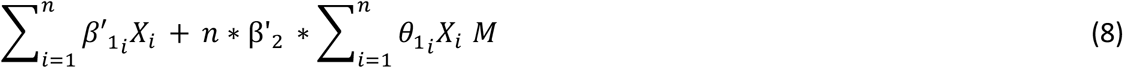

It is important to note that the value of the mediator varies between individuals, the term is the same for the entire population, and therefore a non-informative constant, leaving us with the final formula for the polygenic risk score of direct and indirect effects:

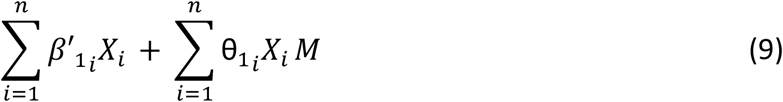

In conclusion, a risk prediction model that integrates clinical and genetic risk factors is defined by the genetic risk score of the direct effect and the genetic risk of the mediator multiplied by the value of mediator M. This model can be expanded to multiple mediator scenario in an additive fashion. The risk model integrating clinical covariates, direct effects, and indirect effects of multiple mediators is defined as follows:

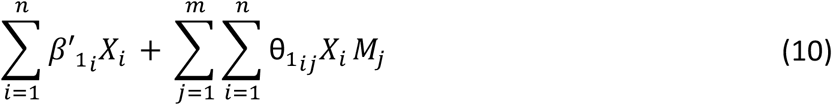

Where still represent the direct effect on SNP on outcome, now adjusted for mediators. It also important to note that (e.g. the effect genetic variant on the mediator) also represents direct effects that has been adjusted for baseline covariates and other mediators.

We emphasize that the choice of GRS should align with its use: if one is predicting cirrhosis risk *in absence of any clinical information*, GRS_total_ is appropriate because it captures all genetic risk. But if one is incorporating genetic risk *alongside clinical mediators*, then GRS_direct_ is more appropriate, to avoid double counting the mediated pathways as described in the Results and shown in Supplementary Table 19.

### Comparing performance of various integrative clinical and genetic risk models

We assessed the performance of the various risk models for cirrhosis by examining metrics such as the AUC (C-statistic) and performing likelihood ratio tests for nested models. Logistic regression was performed using the ‘glm()’ function from “stats” package in R. We used the ROCR R package to compute AUCs and plotted ROC curves for key models. The C-statistic (identical to AUC for binary outcomes) was also computed via the Cstat() function of the DescTools package for confirmation. To compare nested models, we used the likelihood ratio test (lrtest() in the lmtest package) and evaluated differences in AIC, reporting the corresponding chi-square statistics and P-values (Supplementary Table 20).

### Proteome-wide drug target Mendelian randomization for cirrhosis and HCC

We carried out two-sample Mendelian randomization to evaluate whether circulating protein levels have causal effects on risk of cirrhosis and HCC. We used summary-level genetic association from a published plasma proteome GWAS in deCODE study, which measured 4,907 circulating proteins measured using an aptamer-based assays (SOMAscan) in 35,559 Icelanders.^106^ We treated each protein as an exposure and identified cis-acting genetic instruments: genetic variants located within 250kb and of the encoding gene that are associated with the circulating protein (p <5×10^−8^). When multiple such variants existed for a protein, we LD-pruned them (r^2^ <0.01) to retain near-independent instruments using the 1000 Genomes Phase 3 EUR reference panel using the *MendelianRandomization* package in R.^107^. We when extracted the association statistic for those variants from our cirrhosis and HCC multi-ancestry meta-analysis. We aligned effect alleles across datasets and performed Mendelian randomization. When more than one cis-acting genetic variants associated with the protein at genome-wide significance were available within this window, Mendelian randomization was performed using inverse variance weighting. When a single variant within this window was present, Mendelian randomization was performed using the ratio of coefficients method (Supplementary Table 21).

We focused on proteins that passed an FDR threshold of 5% in the MR results for either cirrhosis or HCC. To strengthen causal inference and support the hypothesis that a shared genetic signal influences both circulating protein levels and risk of cirrhosis, we performed colocalization analysis using the coloc Bayesian method with default prior.^108^ A high posterior conditional probability of a shared causal variant (>80%) was considered strong colocalization evidence. Proteins meeting both MR significance and colocalization support were highlighted as high-confidence targets.

### Host genetic modifiers of antiviral treatment response in chronic HCV infection

The study population was stratified into two exposure groups based on the type of antiviral therapy received at the index date: a pegylated interferon (pegIFN)/ribavirin (RBV)-treated cohort and a direct-acting antiviral (DAA)-treated cohort. Antiviral agents were identified using the Anatomical Therapeutic Chemical (ATC) classification system: pegIFN/RBV therapy was defined by ATC codes L03AB10 and L03AB11, and DAA therapy by ATC codes J05AP55 and J05AP57.

For the pegIFN-treated cohort, we identified patients who tested positive for hepatitis C virus (HCV) RNA between January 1, 2000, and December 31, 2010. Inclusion was restricted to patients who initiated pegIFN-based therapy during this period for the first time. The index date was defined as the first date of pegIFN treatment within the VA system. We excluded individuals with a prior diagnosis of cirrhosis, HCC, or liver transplantation before the index date, as well as those with co-infection with hepatitis B virus (HBV) or human immunodeficiency virus (HIV).

For the DAA-treated cohort, patients were included if they tested positive for HCV RNA between January 1, 2013, and December 31, 2021, and initiated DAA therapy during this period. The index date was defined as the first date of DAA treatment. Like the pegIFN cohort, we excluded individuals with prior diagnoses of cirrhosis, HCC, or liver transplantation, as well as those with HBV or HIV co-infection. Another exclusion criterion for the DAA-treated cohort was prior pegIFN exposure within the VA system, to prioritize treatment-naïve individuals.

A total of 1,859 European and 990 African American HCV-infected patients were included in the pegIFN-treated cohort. Among them, 500 European and 242 African American patients developed cirrhosis within 10 years of treatment initiation. In the DAA-treated cohort, 2,045 European and 1,083 African American patients were included, with 101 and 51 individuals, respectively, progressing to cirrhosis during the 10-year follow-up period.

The primary outcome was incident cirrhosis. Each patient was followed for up to 10 years from the index date to assess the incidence of cirrhosis. The primary endpoint was defined as the date of first diagnosis of cirrhosis during the follow-up period, death, or by the end of the study, whichever occurred first. To estimate the relative risk of cirrhosis associated with genetic variation, Cox proportional hazards models were employed, incorporating the competing risk of death adjusted for potential confounders, including age, sex, and alcohol use disorder, and Met-ALT, stratified by ancestry and treatment cohort (pegIFN-treated vs. DAA-treated). Data on viral genotype and sustained viral response were not available. Effect heterogeneity by treatment era and within the respective ancestry was quantified using the Cochrane Q statistic. Results are shown in Supplementary Table 22.

## VA Million Veteran Program: Core Acknowledgements for Publications

### MVP Program OfYice

- Sumitra Muralidhar, Ph.D., Program Director US Department of Veterans Affairs, 810 Vermont Avenue NW, Washington, DC 20420
- Jennifer Moser, Ph.D., Associate Director, Scientific Programs US Department of Veterans Affairs, 810 Vermont Avenue NW, Washington, DC 20420
- Jennifer E. Deen, B.S., Associate Director, Cohort & Public Relations US Department of Veterans Affairs, 810 Vermont Avenue NW, Washington, DC 20420

### MVP Executive Committee

- Co-Chair: Philip S. Tsao, Ph.D. VA Palo Alto Health Care System, 3801 Miranda Avenue, Palo Alto, CA 94304
- Co-Chair: Sumitra Muralidhar, Ph.D. US Department of Veterans Affairs, 810 Vermont Avenue NW, Washington, DC 20420
- J. Michael Gaziano, M.D., M.P.H. VA Boston Healthcare System, 150 S. Huntington Avenue, Boston, MA 02130
- Elizabeth Hauser, Ph.D. Durham VA Medical Center, 508 Fulton Street, Durham, NC 27705
- Amy Kilbourne, Ph.D., M.P.H. VA HSR&D, 2215 Fuller Road, Ann Arbor, MI 48105
- Michael Matheny, M.D., M.S., M.P.H. VA Tennessee Valley Healthcare System, 1310 24th Ave. South, Nashville, TN 37212
- Dave Oslin, M.D. Philadelphia VA Medical Center, 3900 Woodland Avenue, Philadelphia, PA 19104
- Deepak Voora, MD Durham VA Medical Center, 508 Fulton Street, Durham, NC 27705

### MVP Co-Principal Investigators

- J. Michael Gaziano, M.D., M.P.H. VA Boston Healthcare System, 150 S. Huntington Avenue, Boston, MA 02130
- Philip S. Tsao, Ph.D. VA Palo Alto Health Care System, 3801 Miranda Avenue, Palo Alto, CA 94304

### MVP Core Operations

- Jessica V. Brewer, M.P.H., Director, MVP Cohort Operations VA Boston Healthcare System, 150 S. Huntington Avenue, Boston, MA 02130
- Mary T. Brophy M.D., M.P.H., Director, VA Central Biorepository VA Boston Healthcare System, 150 S. Huntington Avenue, Boston, MA 02130
- Kelly Cho, M.P.H, Ph.D., Director, MVP Phenomics VA Boston Healthcare System, 150 S. Huntington Avenue, Boston, MA 02130
- Lori Churby, B.S., Director, MVP Regulatory Affairs VA Palo Alto Health Care System, 3801 Miranda Avenue, Palo Alto, CA 94304
- Scott L. DuVall, Ph.D., Director, VA Informatics and Computing Infrastructure (VINCI) VA Salt Lake City Health Care System, 500 Foothill Drive, Salt Lake City, UT 84148
- Saiju Pyarajan Ph.D., Director, Data and Computational Sciences VA Boston Healthcare System, 150 S. Huntington Avenue, Boston, MA 02130
- Robert Ringer, Pharm.D., Director, VA Albuquerque Central Biorepository New Mexico VA Health Care System, 1501 San Pedro Drive SE, Albuquerque, NM 87108
- Luis E. Selva, Ph.D., Director, MVP Biorepository Coordination VA Boston Healthcare System, 150 S. Huntington Avenue, Boston, MA 02130
- Shahpoor (Alex) Shayan, M.S., Director, MVP PRE Informatics VA Boston Healthcare System, 150 S. Huntington Avenue, Boston, MA 02130
- Brady Stephens, M.S., Principal Investigator, MVP Information Center Canandaigua VA Medical Center, 400 Fort Hill Avenue, Canandaigua, NY 14424
- Stacey B. Whitbourne, Ph.D., Director, MVP Cohort Development and Management VA Boston Healthcare System, 150 S. Huntington Avenue, Boston, MA 02130

### MVP Publications and Presentations Committee

- Co-Chair: Themistocles L. Assimes, M.D., Ph. D VA Palo Alto Health Care System, 3801 Miranda Avenue, Palo Alto, CA 94304
- Co-Chair: Adriana Hung, M.D.; M.P.H VA Tennessee Valley Healthcare System, 1310 24th Ave. South, Nashville, TN 37212
- Co-Chair: Henry Kranzler, M.D. Philadelphia VA Medical Center, 3900 Woodland Avenue, Philadelphia, PA 19104

### DBDS Genomics Consortium

Karina Banasik

Department of Obstetrics and Gynaecology, Copenhagen University Hospital, Hvidovre

Hospital, Copenhagen, Denmark

Jakob Bay

Department of Clinical Immunology, Zealand University Hospital, Køge, Denmark

Andrea Barghetti

Department of Clinical Immunology, Copenhagen University Hospital, Rigshospitalet,

Copenhagen, Denmark

Mette Skou Bendtsen

Department of Clinical Immunology, Copenhagen University Hospital, Rigshospitalet,

Copenhagen, Denmark

Jens Kjærgaard Boldsen

Department of Clinical Immunology, Aarhus University Hospital, Aarhus, Denmark

Søren Brunak

Novo Nordisk Foundation Center for Protein Research, Faculty of Health and Medical

Sciences, University of Copenhagen, Copenhagen, Denmark

Nanna Brøns

Department of Clinical Immunology, Copenhagen University Hospital, Rigshospitalet,

Copenhagen, Denmark

Alfonso Buil Demur

Institute of Biological Psychiatry, Mental Health Centre, Sct. Hans, Copenhagen University

Hospital, Roskilde, Denmark

Johan Skov Bundgaard

Department of Clinical Immunology, Copenhagen University Hospital, Rigshospitalet,

Copenhagen, Denmark

Lea Arregui Nordahl Christoffersen

Department of Clinical Immunology, Zealand University Hospital, Køge, Denmark

Maria Didriksen

Department of Clinical Immunology, Copenhagen University Hospital, Rigshospitalet,

Copenhagen, Denmark

Khoa Manh Dinh

Department of Clinical Immunology, Aarhus University Hospital, Aarhus, Denmark

Joseph Dowsett

Department of Clinical Immunology, Copenhagen University Hospital, Rigshospitalet,

Copenhagen, Denmark

Christian Erikstrup

Department of Clinical Immunology, Aarhus University Hospital, Aarhus, Denmark

Josephine Gladov

Department of Clinical Immunology, Aarhus University Hospital, Aarhus, Denmark

Daniel Gudbjartsson

deCODE Genetics, Reykjavik, Iceland Thomas Folkmann Hansen

Danish Headache Center

Department of Neurology, Copenhagen University Hospital, Rigshospitalet-Glostrup,

Copenhagen, Denmark

Dorte Helenius Mikkelsen

Institute of Biological Psychiatry, Mental Health Centre, Sct. Hans, Copenhagen

University Hospital, Roskilde, Denmark

Lotte Hindhede

Department of Clinical Immunology, Aarhus University Hospital, Aarhus, Denmark

Henrik Hjalgrim

Danish Cancer Society Research Center, Copenhagen, Denmark

Jakob Hjorth von Stemann

Department of Clinical Immunology, Copenhagen University Hospital, Rigshospitalet,

Copenhagen, Denmark

Bitten Aagaard Jensen

Department of Clinical Immunology, Aalborg University Hospital, Aalborg, Denmark

Kathrine Kaspersen

Department of Clinical Immunology, Aarhus University Hospital, Aarhus, Denmark

Bertram Dalskov Kjerulff

Department of Clinical Immunology, Aarhus University Hospital, Aarhus, Denmark

Lisette Kogelman

Danish Headache Center, Department of Neurology, Copenhagen University Hospital, Rigshospitalet-Glostrup, Copenhagen, Denmark

Mette Kongstad,

Department of Clinical Immunology, Copenhagen University Hospital, Rigshospitalet,

Copenhagen, Denmark

Susan Mikkelsen,

Department of Clinical Immunology, Aarhus University Hospital, Aarhus, Denmark

Christina Mikkelsen,

Department of Clinical Immunology, Copenhagen University Hospital, Rigshospitalet,

Copenhagen, Denmark

Line Hjorth Sjernholm Nielsen

Department of Clinical Immunology, Aarhus University Hospital, Aarhus, Denmark

Janna Nissen

Department of Clinical Immunology, Copenhagen University Hospital, Rigshospitalet,

Copenhagen, Denmark

Mette Nyegaard

Department of Health Science and Technology, Faculty of Medicine, Aalborg Univeristy,

Aalborg, Denmark

Sisse Rye Ostrowski

Department of Clinical Immunology, Copenhagen University Hospital, Rigshospitalet,

Copenhagen, Denmark

Frederikke Byron Pedersen

Department of Clinical Immunology, Copenhagen University Hospital, Rigshospitalet,

Copenhagen, Denmark

Ole Birger Pedersen

Department of Clinical Immunology, Zealand University Hospital, Køge, Denmark

Liam James Elgaard Quinn

Department of Clinical Immunology, Zealand University Hospital, Køge, Denmark

Þórunn Rafnar

deCODE Genetics, Reykjavik, Iceland

Palle Duun Rohde

Department of Health Science and Technology, Faculty of Medicine, Aalborg Univeristy,

Aalborg, Denmark

Klaus Rostgaard

Danish Cancer Society Research Center, Copenhagen, Denmark

Andrew Joseph Schork

Institute of Biological Psychiatry, Mental Health Centre, Sct. Hans, Copenhagen University Hospital, Roskilde, Denmark

Michael Schwinn

Department of Clinical Immunology, Copenhagen University Hospital, Rigshospitalet,

Copenhagen, Denmark

Erik Sørensen

Department of Clinical Immunology, Copenhagen University Hospital, Rigshospitalet,

Copenhagen, Denmark

Kari Stefansson

deCODE Genetics, Reykjavik, Iceland

Hreinn Stefánsson

deCODE Genetics, Reykjavik, Iceland

Jacob Træholt

Department of Clinical Immunology, Copenhagen University Hospital, Rigshospitalet,

Copenhagen, Denmark

Unnur Þorsteinsdóttir

deCODE Genetics, Reykjavik, Iceland

Mie Topholm Bruun

Department of Clinical Immunology, Odense University Hospital, Odense, Denmark

Henrik Ullum, Statens Serum Institut, Copenhagen, Denmark

Thomas Werge

Institute of Biological Psychiatry, Mental Health Centre, Sct. Hans, Copenhagen University

Hospital, Roskilde, Denmark

David Westergaard

Department of Obstetrics and Gynaecology, Copenhagen University Hospital, Hvidovre

Hospital, Copenhagen, Denmark

